# Population immunity to pre-Omicron and Omicron SARS-CoV-2 variants in US states and counties through December 1, 2021

**DOI:** 10.1101/2021.12.23.21268272

**Authors:** Fayette Klaassen, Melanie H. Chitwood, Ted Cohen, Virginia E. Pitzer, Marcus Russi, Nicole A. Swartwood, Joshua A. Salomon, Nicolas A. Menzies

## Abstract

Prior infection and vaccination both contribute to population-level SARS-CoV-2 immunity. We used a Bayesian model to synthesize evidence and estimate population immunity to prevalent SARS-CoV-2 variants in the United States over the course of the epidemic until December 1, 2021, and how this changed with the introduction of the Omicron variant. We used daily SARS-CoV-2 infection estimates and vaccination coverage data for each US state and county. We estimated relative rates of vaccination conditional on previous infection status using the Census Bureau’s Household Pulse Survey. We used published evidence on natural and vaccine-induced immunity, including waning and immune escape. The estimated percentage of the US population with a history of SARS-CoV-2 infection or vaccination as of December 1, 2021, was 88.2% (95%CrI: 83.6%-93.5%), compared to 24.9% (95%CrI: 18.5%-34.1%) on January 1, 2021. State-level estimates for December 1, 2021, ranged between 76.9% (95%CrI: 67.6%-87.6%, West Virginia) and 94.4% (95%CrI: 91.2%-97.3%, New Mexico). Accounting for waning and immune escape, the effective protection against the Omicron variant on December 1, 2021, was 21.8% (95%CrI: 20.7%-23.4%) nationally and ranged between 14.4% (95%CrI: 13.2%-15.8%, West Virginia), to 26.4% (95%CrI: 25.3%-27.8%, Colorado). Effective protection against severe disease from Omicron was 61.2% (95%CrI: 59.1%-64.0%) nationally and ranged between 53.0% (95%CrI: 47.3%-60.0%, Vermont) and 65.8% (95%CrI: 64.9%-66.7%, Colorado). While over three-quarters of the US population had prior immunological exposure to SARS-CoV-2 via vaccination or infection on December 1, 2021, only a fifth of the population was estimated to have effective protection to infection with the immune-evading Omicron variant.

**Significance:** Both SARS-CoV-2 infection and COVID-19 vaccination contribute to population-level immunity against SARS-CoV-2. This study estimates the immunity and effective protection against future SARS-CoV-2 infection in each US state and county over 2020-2021. The estimated percentage of the US population with a history of SARS-CoV-2 infection or vaccination as of December 1, 2021, was 88.2% (95%CrI: 83.6%-93.5%). Accounting for waning and immune escape, protection against the Omicron variant was 21.8% (95%CrI: 20.7%-23.4%). Protection against infection with the Omicron variant ranged between 14.4% (95%CrI: 13.2%-15.8%%, West Virginia) and 26.4% (95%CrI: 25.3%-27.8%, Colorado) across US states. The introduction of the immune-evading Omicron variant resulted in an effective absolute increase of approximately 30 percentage points in the fraction of the population susceptible to infection.

## Introduction

As of December 1, 2021, more than 48 million COVID-19 cases and over 780,000 COVID-19-associated deaths had been reported in the United States^1,2^. Between December 1, 2021, and February 1, 2022, an additional 26 million cases (35% of cumulative US COVID-19 cases) and 100,000 deaths (11% of all US COVID-19 deaths) were reported^3^. The epidemic has overloaded healthcare systems, reduced economic activity, and decreased mental health^4-7^. Minimizing COVID-19 morbidity and mortality depends largely on reaching high levels of population immunity. The emergence of the Omicron variant^8,9^ illustrates the importance of identifying areas of highest vulnerability, and underscores how continued evolution of viral variants may reduce effective protection.

The true number of SARS-CoV-2 infections that have occurred since the start of the pandemic is unknown. Recent estimates^10-13^ of the percentage of the US population ever infected vary between 37% and 62%. Seroprevalence estimates from a nationwide convenience sample in commercial laboratories suggested that as of December 21, 2021, 33.5% of the US population over 16 has infection-induced SARS-CoV-2 antibodies^14^, and a nationwide blood donor seroprevalence study estimates 28.8% infection-induced seroprevalence on the same date^15^. The level of protection that infection confers, and the rate at which protection wanes, are incompletely understood^16-18^.

By December 1, 2021, over 240 million US residents (72.9%) had received at least one dose of a COVID-19 vaccine^1^, and over 80 million residents had received both the initial one- or two-dose schedule and a booster. Reported efficacy against symptomatic infection for the three main vaccines available in the US ranged from 66% (Johnson &Johnson) to 94% (Pfizer and Moderna) in clinical trials^19-21^. Vaccine efficacy against infection was estimated to be lower during the Delta surge compared to earlier waves, and initial data indicate further reductions in efficacy against the Omicron variant^8,9^. Declines in vaccine efficacy reflect a possible combination of waning immunity and increased potential for immune escape in viral variants. Despite evidence of waning efficacy^22-25^, vaccination appears to provide durable protection against severe disease, and boosters appear to restore vaccine efficacy toward pre-waning levels.^26-28^

To evaluate the potential for continued SARS-CoV-2 transmission, it is important to estimate overall population immunity to COVID-19. A recent study reported state-level estimates of infection- and vaccine-induced SARS-CoV-2 seroprevalence based on blood donation data^29^, with estimates for May 2021 ranging from 63.7% in Mississippi to 91.7% in Connecticut. While these estimates provide a direct measure of seroprevalence in the study populations, they may be affected by systematic differences between blood donors and the general population. Moreover, these data do not provide county-level estimates or account for waning of protection over time.

For this study we used modeled estimates of cumulative SARS-CoV-2 infections at state- and county-level, and reported coverage for initial and booster vaccination^10,30^. We used survey data to estimate the joint distribution of prior SARS-CoV-2 infection and vaccination^31^. Using these inputs in a Bayesian analytic framework, we estimated the population with SARS-CoV-2 immunological exposure (ever infected or vaccinated) for each US state and county over the course of the epidemic through December 1, 2021. Incorporating evidence on the time-course of natural and vaccine-induced immunity, we estimated effective population immunity against infection and against severe disease over time. We estimated the effective protection against the Omicron variant, accounting for immune escape.

## Results

By December 1, 2021, 59.2% (95%CrI: 46.9%—75.6%) of the US population was estimated to have been infected with SARS-CoV-2, with state-level estimates ranging from 24.0% (95%CrI: 16.0%—40.3%, Hawaii) to 78.5% (95%CrI: 68.7%—88.6%, New Mexico). County-level estimates ranged from 9.0% (San Juan County, Washington) to 91.3% (San Juan County, New Mexico). The percentage of the US population that received at least one COVID-19 vaccine dose was estimated to be 65.2%. State-level coverage varied between 32.0% (West Virginia) and 82.8% (New Hampshire) and county-level coverage varied between 13.3% (Morgan County, West Virginia) and 89.9% (Pitkin County, Colorado).

Based on the results of the Household Pulse Survey, individuals reporting a prior COVID-19 diagnosis were substantially less likely to report being vaccinated. The odds ratio of vaccination among individuals with a prior COVID-19 diagnosis (compared to no prior diagnosis) varied from 0.40 (95%CrI: 0.36–0.44) in Florida to 0.58 (95%CrI: 0.53—0.63) in Texas, with a national mean of 0.52 (95%CrI: 0.50—0.55).

### Immunological exposure

The national estimate for the population *immunologically exposed* was 88.2% (95%CrI: 83.6%— 93.5%). State-level estimates ranged from 76.9% (95%CrI: 67.6%—87.6%, West Virginia) to 94.4% (95%CrI: 91.2%—97.3%, New Mexico; Table 1). Across counties, the percentage *immunologically exposed* ranged from 42.4% (Sioux County, Nebraska) to 98.3% (San Juan County, New Mexico; interquartile range 80.2—87.3%; Figure 1, Figure S2).

**Table 1:** Key population immunity outcomes for each US state on December 1, 2021.

| State | Percentage ever vaccinated* | Percentage ever infected (95% CrI) | Ratio percentage vaccinated / percentage infected | Percentage immunologically exposed (95% CrI) | Percentage effectively protected against infection, pre-Omicron variants (95% CrI) ** | Percentage effectively protected against severe disease, pre-Omicron variants (95% CrI) ** | Percentage effectively protected against infection with Omicron (95% CrI)* ** | Percentage effectively protected against severe disease from Omicron (95% CrI)* ** |
| --- | --- | --- | --- | --- | --- | --- | --- | --- |
| Alabama | 52.4% | 71.3% (60%-84.2%) | 0.73 | 88.6% (83%-94.3%) | 52.8% (50%-55.9%) | 74.1% (70.3%-78.2%) | 20.7% (19.9%-21.7%) | 61% (57.9%-64.3%) |
| Alaska | 58.3% | 66.8% (54.8%-81.2%) | 0.87 | 88.7% (83.3%-94.2%) | 57.7% (54.2%-61.4%) | 73.9% (70.4%-77.9%) | 24.2% (23.2%-25.4%) | 61.6% (58.8%-64.8%) |
| Arizona | 62.5% | 77.6% (67.6%-88.1%) | 0.81 | 93.4% (89.7%-96.8%) | 58.8% (56.1%-61.5%) | 77.3% (75%-79.5%) | 22.9% (22.1%-23.8%) | 63.7% (61.9%-65.5%) |
| Arkansas | 52.8% | 64.3% (52%-79.4%) | 0.82 | 85.8% (79.5%-92.6%) | 50.5% (47.4%-54.5%) | 71.5% (67.3%-76.3%) | 20.6% (19.8%-21.9%) | 59.1% (55.8%-63%) |
| California | 71.4% | 57.2% (44.5%-74.1%) | 1.25 | 89.4% (85.6%-93.9%) | 55.3% (51.3%-60.8%) | 75.3% (73.4%-77.9%) | 22.1% (20.9%-23.8%) | 62.2% (60.7%-64.3%) |
| Colorado | 79.1% | 63.7% (51.4%-79%) | 1.24 | 93.3% (90.6%-96.3%) | 63.2% (59.6%-67.8%) | 78.9% (77.9%-80.1%) | 26.4% (25.3%-27.8%) | 65.8% (64.9%-66.7%) |
| Connecticut | 82.3% | 48.4% (36.1%-66.7%) | 1.7 | 91.6% (89.2%-94.8%) | 58.8% (54.9%-64.8%) | 79.5% (78.7%-80.7%) | 24.1% (23%-25.9%) | 65.9% (65.3%-66.8%) |
| Delaware | 69.4% | 60.3% (47.8%-76.5%) | 1.15 | 89.8% (85.6%-94.5%) | 56.8% (53.2%-61.7%) | 75.8% (73.5%-78.6%) | 23.2% (22.2%-24.7%) | 62.8% (61%-65.1%) |
| District of Columbia | 78.3% | 48.8% (36.5%-67.1%) | 1.6 | 90.0% (86.9%-94%) | 55.7% (51.8%-61.8%) | 77.3% (76.2%-79.1%) | 21.2% (20%-23%) | 63.4% (62.5%-64.8%) |
| Florida | 70.7% | 69% (57.3%-82.6%) | 1.03 | 93.2% (90%-96.4%) | 60.5% (56.7%-64.8%) | 78.5% (76.5%-80.7%) | 23.5% (22.3%-24.8%) | 64.7% (63%-66.5%) |
| Georgia | 50.8% | 71.2% (59.9%-84.1%) | 0.71 | 88.4% (82.8%-94.1%) | 51.8% (48.7%-55.1%) | 73.4% (69.5%-77.6%) | 20.2% (19.2%-21.2%) | 60.3% (57.2%-63.7%) |
| Hawaii | 82.1% | 24% (16%-40.3%) | 3.43 | 86.8% (85.2%-90%) | 46.4% (43.5%-52.6%) | 77.3% (76.9%-78.3%) | 17.8% (16.9%-19.6%) | 63.2% (62.9%-64%) |
| Idaho | 49.4% | 68.6% (56.9%- | 0.72 | 86.8% (80.4%-93.4%) | 54.7% (51.7%- | 72.5% (68.1%- | 23.4% (22.6%- | 60.4% (57%-64.3%) |
|  |  | 82.4%) |  |  | 57.5%) | 77.3%) | 24.3%) |  |
| Illinois | 68.3% | 52.4%<br>(39.9%-70.2%) | 1.3 | 87.2%<br>(82.9%-92.6%) | 52.6%<br>(48.9%-58.1%) | 73.3%<br>(71%-76.6%) | 22.2%<br>(21.1%-23.9%) | 60.9%<br>(59.1%-63.5%) |
| Indiana | 54.9% | 55.2%<br>(42.6%-72.5%) | 1 | 82.9%<br>(76.6%-90.4%) | 47.9%<br>(44.4%-53%) | 68.8%<br>(64.9%-74.2%) | 19.8%<br>(18.8%-21.4%) | 57%<br>(53.8%-61.3%) |
| Iowa | 60.8% | 56.4%<br>(43.8%-73.5%) | 1.08 | 85.3%<br>(79.7%-91.9%) | 53%<br>(50%-57.3%) | 71.4%<br>(68.2%-75.7%) | 23.9%<br>(23.1%-25.2%) | 60%<br>(57.4%-63.4%) |
| Kansas | 61.4% | 59.3%<br>(46.7%-75.7%) | 1.04 | 86.8%<br>(81.4%-92.8%) | 52.6%<br>(49.2%-57.2%) | 72.3%<br>(69.1%-76.2%) | 21.7%<br>(20.7%-23.1%) | 59.9%<br>(57.4%-63.1%) |
| Kentucky | 56.3% | 66.8%<br>(54.8%-81.2%) | 0.84 | 87.8%<br>(82.1%-93.7%) | 55.1%<br>(51.9%-58.6%) | 73.9%<br>(70.1%-78.1%) | 23%<br>(22.1%-24.1%) | 61.4%<br>(58.4%-64.8%) |
| Louisiana | 53.8% | 67.2%<br>(55.2%-81.4%) | 0.8 | 87.2%<br>(81.4%-93.4%) | 51.1%<br>(47.9%-54.9%) | 72%<br>(68.3%-76.3%) | 20.5%<br>(19.6%-21.7%) | 59.5%<br>(56.5%-62.8%) |
| Maine | 78.3% | 36.6%<br>(25.8%-55.3%) | 2.14 | 87.7%<br>(84.7%-92.1%) | 55.5%<br>(52.3%-61.2%) | 77.2%<br>(75.9%-79.4%) | 24.2%<br>(23.3%-25.9%) | 64.4%<br>(63.4%-66.2%) |
| Maryland | 75.8% | 46.5%<br>(34.3%-65%) | 1.63 | 88.6%<br>(85.2%-93%) | 55.1%<br>(51.6%-60.9%) | 75.8%<br>(74.4%-77.9%) | 23.1%<br>(22%-24.8%) | 62.9%<br>(61.8%-64.6%) |
| Massachusetts | 77.7% | 51.7%<br>(39.2%-69.7%) | 1.5 | 90.6%<br>(87.5%-94.5%) | 57.4%<br>(53.5%-63.3%) | 77.9%<br>(76.5%-79.9%) | 23.4%<br>(22.3%-25.2%) | 64.5%<br>(63.4%-66.1%) |
| Michigan | 56.5% | 63.4%<br>(51.1%-78.8%) | 0.89 | 87.5%<br>(81.8%-93.4%) | 53.3%<br>(50.2%-57.1%) | 72.6%<br>(68.9%-77.1%) | 23.2%<br>(22.4%-24.5%) | 60.7%<br>(57.7%-64.2%) |
| Minnesota | 64.8% | 51.7%<br>(39.2%-69.6%) | 1.25 | 85.4%<br>(80.5%-91.6%) | 54.2%<br>(51%-58.8%) | 71.9%<br>(69.2%-75.7%) | 24.5%<br>(23.7%-25.9%) | 60.4%<br>(58.3%-63.5%) |
| Mississippi | 52.8% | 68.3%<br>(56.5%-82.2%) | 0.77 | 87.7%<br>(81.8%-93.7%) | 50.8%<br>(47.7%-54.5%) | 72.4%<br>(68.6%-76.7%) | 20%<br>(19.1%-21.2%) | 59.6%<br>(56.6%-63.1%) |
| Missouri | 54.2% | 51.4%<br>(38.9%-69.3%) | 1.06 | 80.8%<br>(74.3%-89.1%) | 46.8%<br>(43.4%-52%) | 67.8%<br>(63.6%-73.6%) | 19.7%<br>(18.8%-21.3%) | 56.2%<br>(52.9%-60.9%) |
| Montana | 56.6% | 62.9%<br>(50.5%-78.4%) | 0.9 | 86.6%<br>(80.6%-93%) | 55.1%<br>(52%-58.7%) | 72.9%<br>(69.1%-77.6%) | 24.1%<br>(23.2%-25.2%) | 61%<br>(57.9%-64.7%) |
| Nebraska | 55% | 59.8%<br>(42.8%-79.1%) | 0.92 | 84.6%<br>(76.2%-92.8%) | 51.8%<br>(46.9%-57.8%) | 70.3%<br>(64.9%-76.2%) | 22.4%<br>(21.1%-24.3%) | 58.7%<br>(54.4%-63.4%) |
| Nevada | 62.3% | 67.7%<br>(49.2%-86.5%) | 0.92 | 90.1%<br>(82.9%-96.3%) | 55.4%<br>(49.2%-62.1%) | 75%<br>(70.4%-79.6%) | 21.5%<br>(19.7%-23.6%) | 61.7%<br>(58%-65.5%) |
| New Hampshire | 82.8% | 39.9%<br>(28.7%-58.7%) | 2.08 | 90.5%<br>(88.3%-93.8%) | 54.9%<br>(50.9%-61.8%) | 80.2%<br>(79.5%-81.4%) | 18.1%<br>(16.9%-20.1%) | 64.7%<br>(64.1%-65.7%) |
| New Jersey | 75.3% | 58.3%<br>(45.7%- | 1.29 | 91.1%<br>(87.8%-95%) | 57.6%<br>(53.6%- | 76.8%<br>(75.3%- | 22.8%<br>(21.6%- | 63.4%<br>(62.2%- |
|  |  | 74.9%) |  |  | 63%) | 78.7%) | 24.5%) | 64.9%) |
| New Mexico | 67.2% | 78.5%<br>(68.7%-88.6%) | 0.86 | 94.4%<br>(91.2%-97.3%) | 62.4%<br>(60%-64.7%) | 78.4%<br>(76.6%-80.1%) | 25.9%<br>(25.2%-26.7%) | 65.2%<br>(63.8%-66.6%) |
| New York | 75.8% | 60.6%<br>(48%-76.7%) | 1.25 | 91.8%<br>(88.7%-95.4%) | 57.8%<br>(53.5%-63.5%) | 77.4%<br>(76%-79.2%) | 21.4%<br>(20.1%-23.2%) | 63.4%<br>(62.2%-64.8%) |
| North Carolina | 58.1% | 51.7%<br>(37.6%-71.7%) | 1.12 | 83.2%<br>(76.5%-91.2%) | 46.2%<br>(41.1%-53.9%) | 69.4%<br>(65.1%-75.4%) | 17.3%<br>(15.8%-19.6%) | 56.7%<br>(53.3%-61.5%) |
| North Dakota | 53.1% | 70%<br>(58.4%-83.3%) | 0.76 | 88.4%<br>(82.6%-94.2%) | 53.2%<br>(50.6%-56%) | 72.6%<br>(68.9%-76.6%) | 22.8%<br>(22.1%-23.8%) | 60.5%<br>(57.6%-63.7%) |
| Ohio | 56% | 50.3%<br>(38%-70.7%) | 1.11 | 81.3%<br>(75.2%-90%) | 48.3%<br>(44.9%-54.7%) | 68.2%<br>(64.3%-74.5%) | 21.3%<br>(20.3%-23.2%) | 56.9%<br>(53.8%-62%) |
| Oklahoma | 58.7% | 74.6%<br>(63.8%-86.3%) | 0.79 | 91.4%<br>(86.9%-95.8%) | 57.8%<br>(55%-60.5%) | 76%<br>(73.1%-79%) | 22.6%<br>(21.8%-23.5%) | 62.7%<br>(60.3%-65.1%) |
| Oregon | 69.6% | 42.4%<br>(29.7%-62.9%) | 1.64 | 85.0%<br>(80.4%-91.3%) | 52.8%<br>(48.7%-59.9%) | 73.1%<br>(70.6%-77.1%) | 22.4%<br>(21.3%-24.5%) | 60.8%<br>(58.8%-64%) |
| Pennsylvania | 76.1% | 54.2%<br>(41.6%-71.7%) | 1.4 | 90.6%<br>(87.4%-94.5%) | 56.3%<br>(51.9%-62.5%) | 77.3%<br>(75.7%-79.4%) | 20.7%<br>(19.4%-22.6%) | 63.2%<br>(62%-64.9%) |
| Rhode Island | 75.4% | 64.1%<br>(51.8%-79.2%) | 1.18 | 92.6%<br>(89.3%-96.1%) | 61.2%<br>(57.8%-65.7%) | 78.7%<br>(77.1%-80.6%) | 25.2%<br>(24.2%-26.6%) | 65.3%<br>(64.1%-66.9%) |
| South Carolina | 56.4% | 62.3%<br>(49.9%-78%) | 0.91 | 86.3%<br>(80.3%-92.8%) | 51.9%<br>(48.5%-56.2%) | 72.2%<br>(68.4%-76.8%) | 21%<br>(20%-22.3%) | 59.7%<br>(56.7%-63.4%) |
| South Dakota | 63.5% | 63.6%<br>(51.3%-78.9%) | 1 | 88.8%<br>(83.9%-94.1%) | 55.4%<br>(52.6%-59.1%) | 73.3%<br>(70.7%-76.5%) | 22.9%<br>(22.1%-24.1%) | 60.9%<br>(58.8%-63.4%) |
| Tennessee | 53.8% | 65.2%<br>(51.3%-85.3%) | 0.82 | 86.6%<br>(79.9%-94.9%) | 51.7%<br>(47.6%-58.2%) | 72.3%<br>(67.6%-78.9%) | 21.6%<br>(20.4%-23.7%) | 60%<br>(56.3%-65.4%) |
| Texas | 62.6% | 66.9%<br>(54.9%-81.2%) | 0.94 | 89.4%<br>(84.9%-94.3%) | 54.4%<br>(50.9%-58.6%) | 73.6%<br>(71%-76.7%) | 21.1%<br>(20.1%-22.4%) | 60.6%<br>(58.5%-63%) |
| Utah | 50.5% | 61.5%<br>(49.1%-77.4%) | 0.82 | 83.7%<br>(77.1%-91.2%) | 49%<br>(45.3%-53.3%) | 68.8%<br>(64.5%-74%) | 19.7%<br>(18.6%-21%) | 56.8%<br>(53.4%-61%) |
| Vermont | 61.4% | 35.3%<br>(25.8%-52.7%) | 1.74 | 78%<br>(73.2%-85.6%) | 50.4%<br>(47.6%-55.8%) | 67.8%<br>(64.9%-72.9%) | 24.2%<br>(23.5%-25.7%) | 57.4%<br>(55.2%-61.4%) |
| Virginia | 64.9% | 44.7%<br>(32.7%-63.4%) | 1.45 | 83%<br>(78.1%-89.7%) | 50.1%<br>(46.6%-55.7%) | 70.7%<br>(67.9%-74.9%) | 21.2%<br>(20.3%-22.9%) | 58.7%<br>(56.6%-62.1%) |
| Washington | 69.2% | 41.7%<br>(30.1%-60.5%) | 1.66 | 84.5%<br>(80.2%-90.5%) | 50.9%<br>(47.3%-57%) | 72.3%<br>(70.1%-75.7%) | 21.6%<br>(20.6%-23.3%) | 60%<br>(58.3%-62.8%) |
| West Virginia | 32% | 61.3%<br>(48.8%- | 0.52 | 76.9%<br>(67.6%-87.6%) | 41.6%<br>(37.6%- | 65.4%<br>(58.3%- | 14.4%<br>(13.2%- | 53%<br>(47.3%-60%) |
|  |  | 77.2%) |  |  | 46.2%) | 74.2%) | 15.8%) |  |
| Wisconsin | 63.4% | 51.4%<br>(39.6%-71%) | 1.23 | 84.7%<br>(79.7%-91.7%) | 52.9%<br>(49.7%-58.6%) | 71.7%<br>(68.8%-76.4%) | 23.5%<br>(22.7%-25.3%) | 60.1%<br>(57.8%-63.8%) |
| Wyoming | 51.7% | 72.3%<br>(55.3%-91.4%) | 0.71 | 89.1%<br>(80.5%-97.1%) | 56.2%<br>(50.7%-62.6%) | 74.7%<br>(68.5%-81.3%) | 23.7%<br>(22.1%-25.8%) | 62.1%<br>(57.2%-67.5%) |

\*Received at least one COVID-19 vaccine dose
\*\* Assuming the base-case waning functions.
\*\*\* Assuming the base-case waning functions and the medium immune evasion scenario.

**Figure 1:**
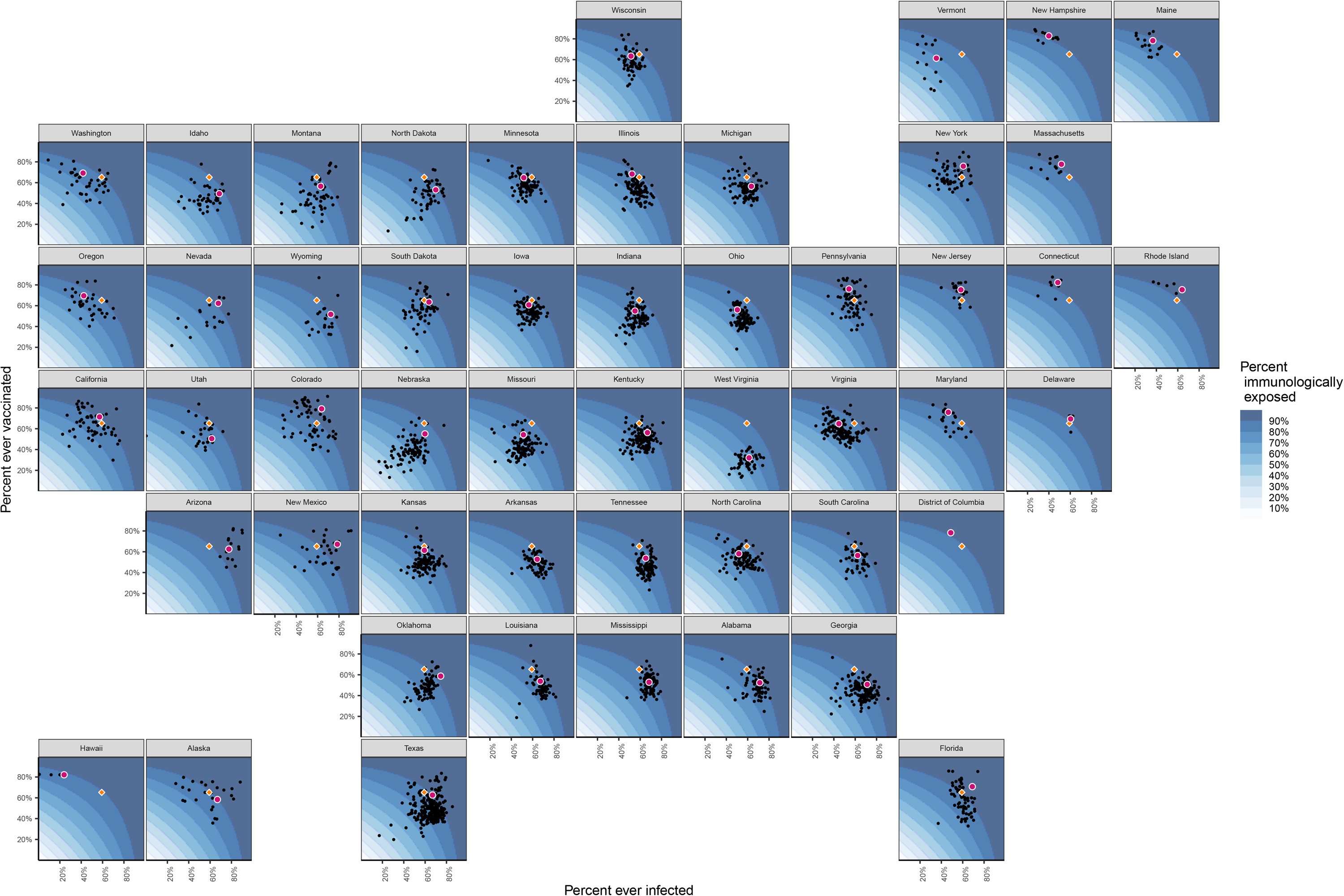
Estimated percentage *immunologically exposed* on December 1, 2021, for each US county and state. Footnote: The background coloring indicates the state-specific distribution of immunity as a function of infections and vaccinations. Black dots represent counties in a state, red dots the state average. The orange diamond represents the state-population-weighted national averages of the percentage ever infected and vaccinated (this does not represent the national average of immunity because the calculation for immunity is state-specific).

### Effective protection

Accounting for waning of immunity, the percentage of the US population with *effective protection* against infection with pre-Omicron variants increased from 14.7% (95%CrI: 11.1%— 19.8%) on January 1, 2021, to 54.1% (95%CrI: 50.5%—59.2%) by December 1, 2021. On this date *effective protection* against infection with the Omicron variant was estimated to be 21.8% (95%CrI: 20.7%—23.4%). The percentage of the population with *effective protection* against severe disease was estimated to be 74.1% (95%CrI: 71.4%—77.6%) for pre-Omicron variants, as opposed to 61.2% (95%CrI: 59.1%—64.0%) for the Omicron variant.

*Effective protection* against infection with Omicron varied at the state level between 14.4% (95%CrI: 13.2%—15.8%, West Virginia) and 26.4% (95%CrI: 25.3%—27.8%, Colorado) and *effective protection* against severe disease ranged between 53.0% (95%Cri: 47.3%—60.0%, West Virginia) and 65.8% (95%CrI: 64.9%—66.7%, Colorado) (Table 1).

Figure 2 shows how the state-level percentage *immunologically exposed, effectively protected* against infection, and *effectively protected* against severe disease has evolved over the course of the epidemic and with the introduction of Omicron. For counties, the percentage of the population with *effective protection* against infection and severe disease caused by pre-Omicron variants, respectively, varied between 26.6% and 48.4% (Cameron Parish, Louisiana) and 72.5% and 77.4% (Fairfax City, Virginia; Figure 3). Estimates of the *effective protection* against infection and severe disease from the Omicron variant varied between 8.4% and 26.4% (McPherson County, Nebraska), and 33.1% and 71.3% (Mineral County, Colorado; Figure S3).

**Figure 2:**
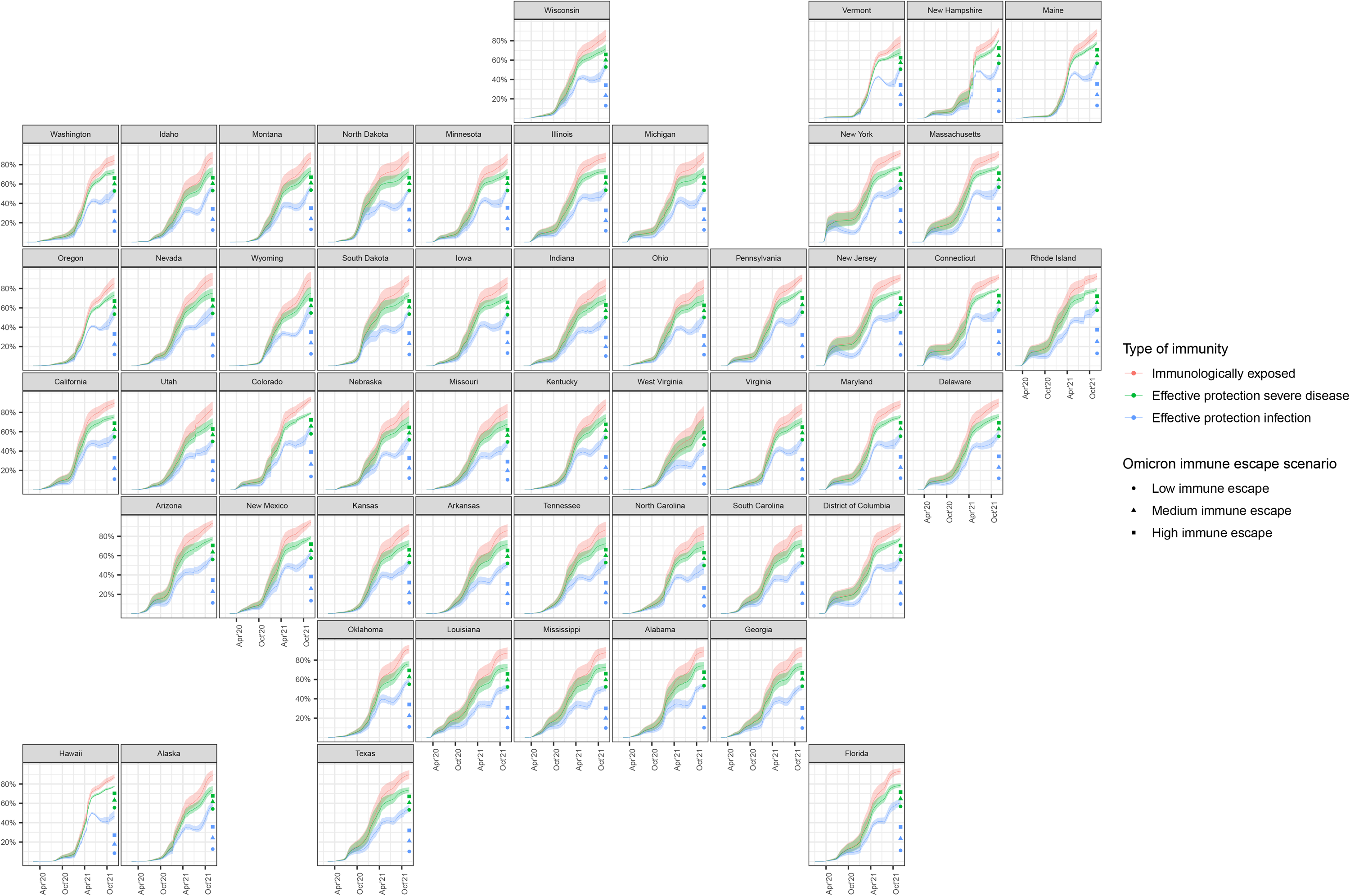
State-level estimates and uncertainty intervals of the percentage *immunologically exposed, effectively protected* against infection, and *effectively protected* against severe disease over time, with estimates of *effective protection* against Omicron infection under three immune escape scenarios.

**Figure 3:**
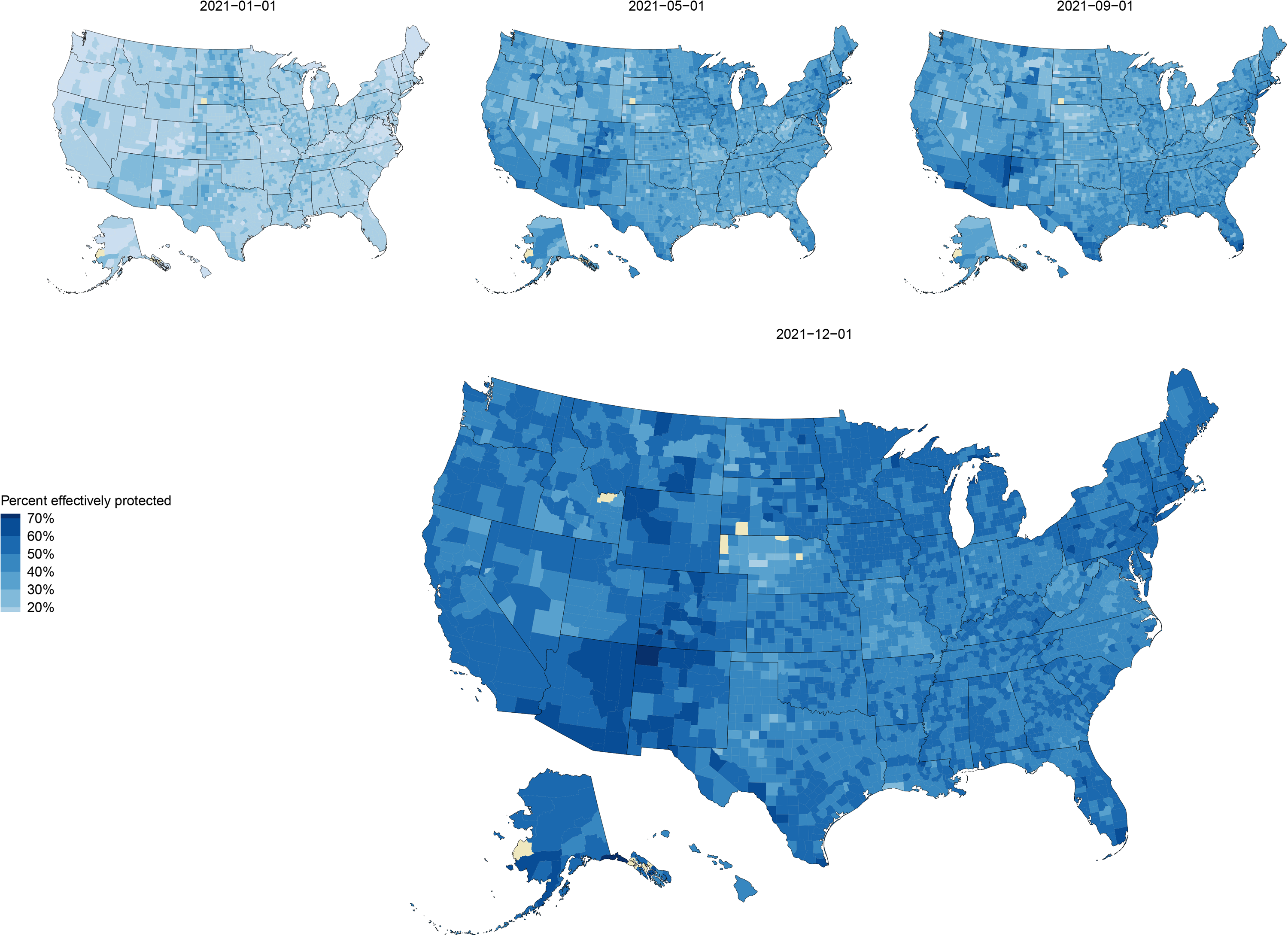
County-level estimates of the percentage of the population *effectively protected* against infection at four time-points between January 31, 2021 and December 1, 2021.

### Relative contributions of prior infection and vaccination

On December 1, 2021, 23.0% (95%CrI: 18.4%—28.3%) of the US population was estimated to have been infected but not vaccinated, 29.0% (95%CrI: 18.0%—36.8%) was estimated to have been vaccinated but not infected, and 36.2% (95%CrI: 28.4%—47.2%) was estimated to have been both vaccinated and infected. The relative contributions of vaccination and prior infection varied widely across states and counties, and over time (Figures S4 and S5). Figure 4 reports the relative contributions of vaccination and prior infection to overall immunity on December 1, 2021, highlighting regional patterns in the different pathways to immunity. The state-population weighted regional averages of the population *immunologically exposed* were 89.1% in the West (20.4% only infected, 30.8% only vaccinated), 84.7% in the Midwest (24.8% only infected, 30.0% only vaccinated), 88.3% in the South (27.5% only infected, 24.4% only vaccinated), and 91.0% in the Northeast (14.5% only infected, 35.5% only vaccinated).

**Figure 4:**
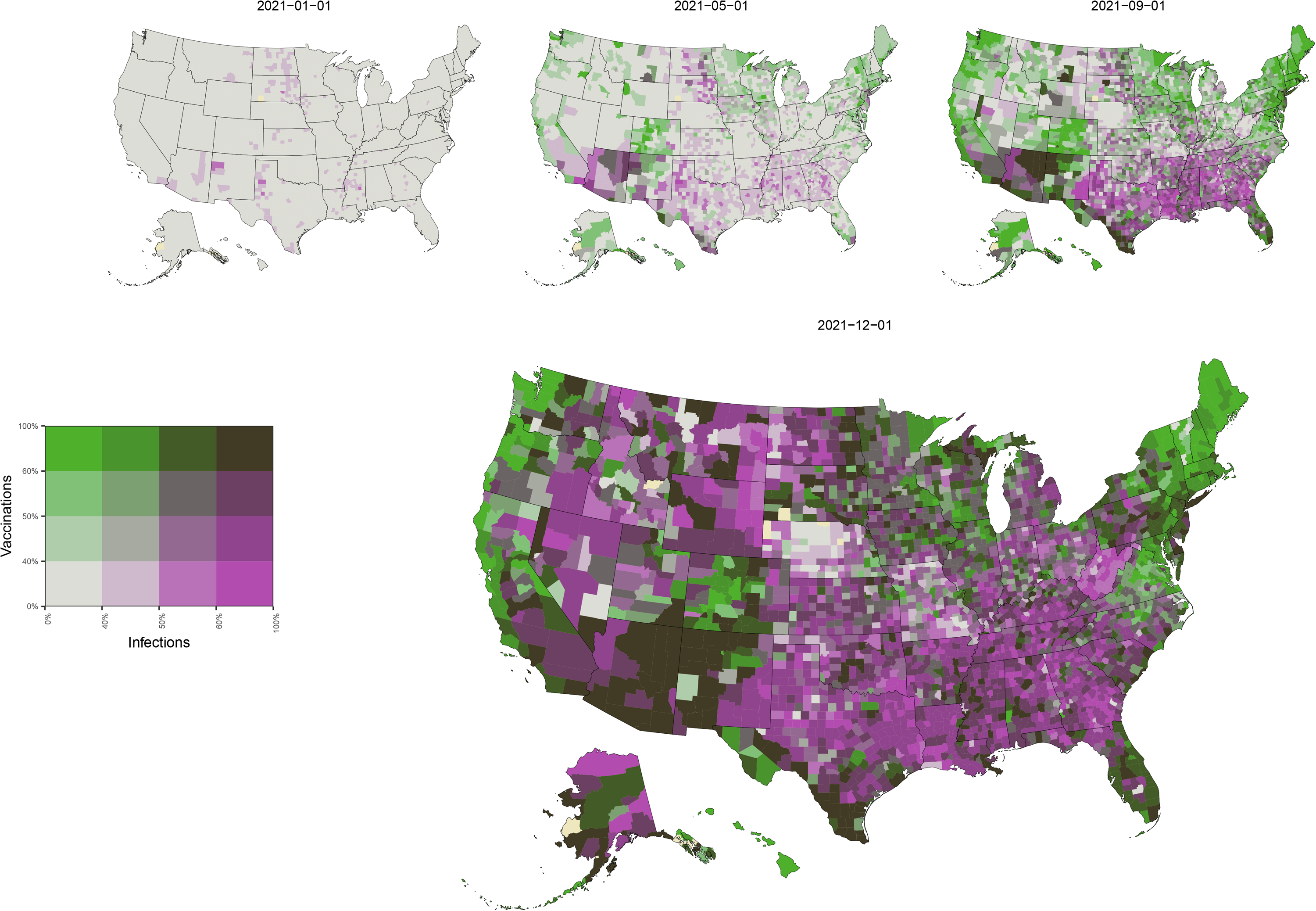
Relative contribution of prior infection and vaccination to population immunity for each county at four time-points between January 31, 2021 and December 1, 2021. Footnote: The percentage ever infected and the percentage vaccinated are categorized with cut-off scores of 40%, 50% and 60%. These values which roughly corresponds to the quantile breakpoints of the estimates of ever infected and vaccinated on December 1, 2021.

### Validation

We re-estimated the odds ratio of vaccination given prior infection using independent survey data, allowing bi-weekly national estimates between January and June 2021^32^ (SI Methods). For the 13 survey waves included in this period, the odds ratio of vaccination given prior infection varied between 0.35 and 0.98, and the mean odds ratio was 0.51 (95%CrI: 0.44—0.59), similar to the estimated national value in the main analysis (0.52; 95%CrI: 0.50—0.55). We compared our estimates of the percentage of the population *immunologically exposed* with blood donor seroprevalence estimates (Figure S6). Our estimates of the percentage *immunologically exposed* were generally lower than seroprevalence estimates from Jones et al.^29^

### Sensitivity analyses

We conducted additional analyses evaluating the sensitivity of our *effectively protected* estimates to waning assumptions, with pessimistic and optimistic scenarios (SI Methods and Figures S1), in combination with the Omicron immune escape scenarios (Table S2). In these analyses national estimates of *effective protection* against infection from pre-Omicron variants ranged between 47.1% and 64.3%, and protection against infection with Omicron ranged between 12.3% and 28.4% (Table S3). *Effective protection* against severe disease from pre-Omicron variants ranged between 67.5% and 79.0%, and protection against severe disease from the Omicron variant ranged between 57.4% and 61.7% (Table S4).

## Discussion

We analyzed the joint distribution of COVID-19 vaccination and prior SARS-CoV-2 infection in each US state and county since the beginning of the COVID-19 epidemic and estimated how population immunity changed over this period. By December 1, 2021, over three-quarters of the US population had prior immunological exposure to SARS-CoV-2 via vaccination or infection, and half of the population retained *effective protection* against infection with previously circulating variants, while only a fifth of the population had *effective protection* against infection with the Omicron variant. There were differences in the overall level of immunity achieved, and the relative contribution of vaccination and prior infection to reaching estimated immunity levels, between states and counties.

To estimate *effective protection*, we made assumptions about how natural and vaccine-induced immunity wanes over time. Despite accumulating evidence these assumptions are still uncertain. In sensitivity analyses, we examined additional waning scenarios, providing a range of possible values for the level of *effective protection*. Recent studies suggest that antibody titers decay rapidly in the three months following infection and more gradually thereafter^18,33^. However, neutralizing antibody activity has been observed up to eight months after symptom onset^16^, and simulation studies suggest that titers wane below 1:20 (often used to infer 50% protection) for the majority of previously-infected individuals by 341 days after symptom onset^18^. Antibody titers in vaccinated individuals are believed to wane at similar rates^17^, although vaccine efficacy has been shown to remain robust in the first six months following inoculation^24,25^. The longer-term effects of waning antibody titers on vaccine efficacy, and the amount of protection conferred by partial vaccination (e.g., one dose of a two-dose regimen) are still incompletely understood. Our assumptions regarding the immune escape of the Omicron variant are preliminary, but we present a range of plausible scenarios, each of which demonstrates the ability for SARS-CoV-2 variants that evade immune protection to spread widely despite high prevalence of prior infection and vaccine coverage.

Our modeled estimates rely on multiple publicly available data sources, each of which has its own limitations. For vaccination coverage, we used data from Merritt *et al*^34,35^, which endeavors to address known biases in CDC vaccination data. We further adjusted these vaccination data to assure that no greater than 100% of the over 12 population could have been vaccinated by the end of our study period. While most indicators suggest lower cumulative infections among children, there is evidence that contradicts this and suggests a higher seroprevalence for children compared to adults^36-39^. For our infection rate estimates, we used a model which leverages case notification and COVID-19 mortality data from Johns Hopkins University; we incorporated modeled uncertainty in these estimates into our model of immunity. The model for infections assumes individuals can only be infected once, so possible reinfections and breakthrough infections amongst vaccinated individuals are not accounted for in our estimates of immunity. Because Omicron breakthrough infections are likely rendering unreliable infection estimates, we only used estimates of infections up until December 1, 2021.

We estimated the relationship between prior infection and vaccination status using survey data that have been criticized for non-representativeness^40^. While this relationship was confirmed in independent survey data that have been validated against external benchmarks^40^, it is still possible that reporting biases could have distorted this relationship. If there is greater overlap between vaccinated and previously infected populations, then overall population immunity will be lower than estimated in our analyses.

The COVID-19 epidemic, measures, and knowledge are changing fast. On November 1, 2021, children 5-11 years old became eligible for vaccination; however, we assume this age group to be unvaccinated throughout the analysis. Any vaccinations of children will count towards the adult immunity estimates. We also did not account for differences in waning depending on the SARS-CoV-2 variant distribution.

## Conclusions

The fraction of the US population that on December 1, 2021, has ever been infected with SARS-CoV-2 and/or received at least one dose of a COVID-19 vaccine varied between counties and states. Accounting for waning of population immunity, *effective protection* against infection by pre-Omicron variants in US states was between 27.6 and 40.4 percentage points lower than the percentage *immunologically exposed*. Introduction and takeover of the Omicron variant reduced *effective protection* against infection by another 26.2 to 37.0 percentage points across US states.

## Methods

### Data

#### Infections

We extracted time-series estimates of SARS-CoV-2 infections from a statistical model^10^ that synthesizes reported data on COVID-19 cases and deaths^3,41^, accounting for under-ascertainment and time lags associated with these reported outcomes. We imputed missing cases and deaths data for Nebraska counties after June 30, 2021 (see SI Methods).

We estimated cumulative infections for each US state and 3137 counties, starting from the first reported case date in each location (range: December 1, 2019 – November 12, 2020). We excluded 6 counties due to missing/insufficient data. As estimates of infection rates during the Omicron wave are highly uncertain, we excluded data after December 1, 2021.

#### Vaccinations

We extracted estimates from a repository reporting weekly county-level vaccination coverage based on CDC-reported data, adjusted for known biases and incompleteness in several states^34,35^. We imputing missing data and smoothed the weekly time-series^42^ into a daily time-series of residents having received at least one vaccine dose (see SI Methods). We summed these counts for all counties within each state to produce state-level estimates. We extracted daily state- and county-level booster coverage data from CDC reports^30^. County-level booster coverage reporting starts on December 16, 2021. Booster coverages before this date were imputed proportional to the corresponding state coverage using the ratio of county to state booster coverage on December 16, 2021.

#### Co-occurrence of infection and vaccination

The Census Bureau’s Household Pulse Survey collects data on COVID-19-relevant beliefs and behaviors at two-weekly intervals, for individuals 18 years and older^31^. We extracted data from February 2 to August 30, 2021, to estimate the joint distribution of infection and vaccination among survey respondents. We extracted the variables *had covid* (Yes/No; whether a respondent has received a positive COVID-19 diagnosis), *received vaccine* (Yes/No; whether a respondent has received as least one dose of a COVID-19 vaccine), *state* and *week*. Responses other than Yes or No (e.g., Unknown) were excluded (2.3% of respondents).

## Estimation

For each location, we computed the percentage *immunologically exposed*, defined as the percentage of the population with a prior SARS-CoV-2 infection, at least one dose of a COVID-19 vaccine, or both. We calculated values separately for individuals aged less than 12 years and those aged 12 years or older, due to differences in vaccine eligibility for these groups over the study period.

### Immunological exposure for the population aged 12 and over

Using the Household Pulse data, we fit a logistic regression model to estimate the association between self-reported vaccination status and prior COVID-19 diagnosis. We operationalized this relationship as the odds ratio for reported vaccination, comparing individuals reporting a prior COVID-19 diagnosis to those reporting no prior diagnosis (see SI Methods). Using these regression results, we created state-specific prior distributions for the odds ratio of vaccination given prior infection status, for individuals aged 12 and older (Table S1). This approach assumes that the odds ratio for vaccination among those with a prior undiagnosed infection is the same as for those with a prior diagnosed infection. We validated this relationship using data from the Axios-Ipsos Coronavirus Tracker^40^.

We calculated the joint probability of being vaccinated or infected as the sum of the marginal probabilities for prior infection and prior vaccination, minus the probability of being both infected and vaccinated, to avoid double counting (see SI Methods).

### Immunological exposure for the population under 12 years old

For the population under 12 years old, the percentage *immunologically exposed* was assumed equal to the estimated percentage ever infected, as this age group was not eligible for vaccination during most of the study period. We assumed infection prevalence in this age group was equal to prevalence in the overall population. We combined under-12 and over-12 immunity estimates in a weighted sum to obtain the percentage *immunologically exposed* in the full population. We validated our results by comparing them to published population immunity estimates based on laboratory data from a blood-donor sample^29^.

### Waning of protection

Protection conferred by natural infection and vaccination appears to decline over time^28,43-45^. We translated the latest available evidence into approximate waning curves under three scenarios: a base-case scenario (used in the main analysis), as well as pessimistic and optimistic scenarios (used in sensitivity analyses). Figure S1 shows assumed levels of protection for each scenario. For the base-case scenario, we assumed that infection or vaccination each initially confer 80% protection against infection that declines to 25% by 12 months after exposure, and protection against severe disease starts at 95% and declines to 85% after 12 months. For individuals both infected and vaccinated, we assumed constant protection of 90% against infection and 95% against severe disease (see SI Methods for optimistic and pessimistic scenarios). We assumed that booster uptake was randomly distributed in the eligible population, and that receiving a booster restored immunity to original (pre-waning) levels and subsequently waned following the ‘both infected and vaccinated’ curve^27,28,46^. Using these assumptions we calculated the percentage *effectively protected* against infection and against severe disease.

### Immune escape under the Omicron variant

Early evidence indicates the Omicron variant can escape immunity acquired by immunization or infection with earlier variants. From the latest available evidence^8,26,47,48^, we constructed *high, medium* (used in the main analysis) and *low* immune-escape scenarios. In the *medium* escape scenario, protection against infection was reduced by 70% (40% for those who received a booster) compared to immunity against pre-Omicron variants. Protection against severe disease reduced by 20% (15% for those who received a booster) compared to immunity against pre-Omicron variants (see Table S2 for *low* and *high* escape scenarios).

### Model implementation

We executed the analysis in R^49^ and the rstan package^50^ (https://github.com/covidestim/covidestim/tree/immunity-waning). For state-level results, we report uncertainty using equal-tailed 95% credible intervals (95%CrI). We calculated national estimates and conservative uncertainty intervals by summing state-level estimates and upper and lower bounds of state-level intervals. County-level estimates were produced using an optimization routine^10^ that produces point estimates without uncertainty intervals. In summarizing county-level results, we excluded counties with a population under 1,000 (0.9% of all counties). All reported results are for December 1, 2021, unless otherwise noted.

## Supporting information

eMaterials

## Data Availability

All data used in the main analysis are available from The Covid Tracking Project, Johns Hopkins CSSE, Github, US Census Bureau, Ipsos and a JAMA publication.
Code for analyses is available from GitHub.
https://covidtracking.com/
https://www.mass.gov/info-details/covid-19-response-reporting
https://www.openicpsr.org/openicpsr/project/144903/version/V1/view
https://github.com/bansallab/vaccinetracking/tree/main/vacc_data
https://www.census.gov/programs-surveys/household-pulse-survey/data.html
https://github.com/covidestim/covidestim/immunity-waning
https://jamanetwork.com/journals/jama/fullarticle/2784013

https://www.mass.gov/info-details/covid-19-response-reporting

https://www.openicpsr.org/openicpsr/project/144903/version/V1/view

https://github.com/bansallab/vaccinetracking/tree/main/vacc_data

https://www.census.gov/programs-surveys/household-pulse-survey/data.html

https://github.com/covidestim/covidestim/immunity-waning

https://jamanetwork.com/journals/jama/fullarticle/2784013

