## Supplementary material for "Population immunity to pre-Omicron and Omicron SARS-CoV-2 variants in US states and counties through December 1, 2021": eMaterials

This file contains:

**SI Methods.**

**Table S1.**

**Table S2.**

**Table S3.**

**Table S4.**

**Figure S1.**

**Figure S2.**

**Figure S3.**

**Figure S4.**

**Figure S5.**

**Figure S6.**

### SI Methods

#### Imputation of cases and deaths data for Nebraska counties

The time-series of cases and deaths data are truncated for counties in Nebraska after June 30, 2021, due to non-reporting of cases and deaths data at the county level. To produce estimates for Nebraska we imputed cases and deaths for each county based on state-level cases and deaths, assuming each county experienced the same proportional changes in cumulative cases and deaths as reported at the state level.

#### Imputation and temporal disaggregation of vaccination data

We extracted estimates from a repository reporting county-level vaccination coverage based on CDC vaccination data, adjusted for known biases and incompleteness in several states[1,2].These estimates include weekly values for the number of county residents having received at least one COVID-19 vaccine dose (*First dose*) and the number of residents having completed an FDA-approved vaccine sequence (*Fully vaccinated*). We imputed missing *First dose* estimates using the observed *Fully vaccinated* data, and smoothed weekly data to create a daily time-series.

We imputed data for 2,006 counties in the weekly vaccination data (*First dose*) *(*minimum missing weeks: 1, 567 counties, maximum missing weeks: 35, 8 counties). The states with the highest percentage of counties with missing data are Nebraska and South Dakota. After the temporal disaggregation of the weekly time series to daily data, a remaining 57 counties had missing data at the end of the time-series (minimum missing days: 8, maximum missing days: 211).

We used a three-step process to render a daily timeseries from the (irregular) weekly timeseries. First, we used linear interpolation to impute missing *First dose* and *Fully vaccinated* data on a weekly (7-day) interval from January 10th, 2021. Second, we disaggregated the weekly time series to the daily level, using a second order smoothness constraint. This method fits a smooth line that always passes the observation on the first day of the week and increases in the following days[3]. Third, we imputed the remaining missing data of at the end of the time series using the following approach.

We imputed the missing data of *First dose* at time X using the fraction of people who get fully vaccinated. We assume there is an average lag of Y days between *First dose* and *Fully vaccinated*. For a single-dose vaccine (Johnson, 8% of all fully vaccinated individuals), the status *Fully vaccinated* is achieved on the same day as the *First dose*. For Moderna (37.3% of all fully vaccinated individuals), the recommended time between the first and second dose is four weeks (28 days). For Pfizer-BioNTech (54.6%), the recommended time is three weeks (21 days) [4,5]. The weighted average of time until fully vaccinated is 22 days.

For each time point, we computed the fraction fully vaccinated. The missing values of *First dose* were calculated from the imputed values and had to meet the constraint that at any time, the total number *First dose* must be smaller than the total population size. We further constrain the time series to be monotonically increasing.

Children under 12 years old were not eligible for COVID-19 vaccination in the time period of analysis. We constrained the fraction of the full population with a *First dose* to never exceed the fraction of the full population of 12 years and older. For example, Santa Cruz, Arizona, reports *99%* of its population has received a *First dose,* while only 81% of the population is 12 years and older. In this case, we constrain the constrain the fraction vaccinated to be 81%.

#### Logistic regression rendering state specific odds ratios of vaccination given infection status

We used a logistic regression model to estimate the association between self-reported vaccination status and prior COVID-19 diagnosis. The regression model included fixed effects for week and state. We also included state-level random coefficients for prior COVID-19 diagnosis, to allow for state-level differences in the overlap between vaccination and prior infection. Below is the full regression model, where $X_{(.)}$ is a model matrix for the specified variable. We used a binomial likelihood to model the number people with a self-reported vaccination status ($n$) out of the total ($N$).

$$\mathrm{logit}(\theta) = \mathrm{b}_0 + X_{\text{week}}\mathbf{b}_{\text{week}} + X_{\text{state}}\mathbf{b}_{\text{state}} + X_{\text{state}\times\text{covid}}(\mathrm{b}_{\text{covid}} + \mathbf{g}_{\text{r.e.}})$$
$$\mathcal{L}(\theta|N, \mathrm{n}) = \theta^n(1-\theta)^{N-n}$$
$$g_{\text{r.e.}} \sim \mathcal{N}(0, \sigma)$$
$$\sigma \sim U(0, \infty)$$

**Derivation of joint probability from odds ratio**

We have estimates of the probability of ever being infected and (adjusted) data of the probability of ever being vaccinated. In addition, we have a specified a data-driven prior distribution on the odds ratio of vaccination for those with a prior diagnosis versus those without a prior diagnosis. We want to compute the probability of being immune (the probability of ever being vaccinated and/or infected).

Following the rules of probability and our definition of immunity, we can compute the probability of being immune as a function of the probability of vaccinated, infected, and the conditional probability of being vaccinated given prior infection.

$$\begin{aligned}
p_{\text{immune}} &= p_{\text{vac}\,\cup\,\text{inf}} \\
&= p_{\text{vac}} + p_{\text{inf}} - p_{\text{vac}\,\cap\,\text{inf}} \\
&= p_{\text{vac}} + p_{\text{inf}} - p_{\text{vac}}\ p_{\text{vac}\,|\,\text{inf}} \\
&= p_{\text{vac}} + p_{\text{inf}} - p_{\text{vac}} \frac{O_{\text{vac}\,|\,\text{inf}}}{1 + O_{\text{vac}\,|\,\text{inf}}}
\end{aligned}$$

To compute the probability of being immune, we need an expression for the unknown $O_{\text{vac}\,|\,\text{inf}}$. Making use of the fact that:

$$\begin{aligned}
\frac{O_{\text{vac}\,|\,\text{inf}}}{O_{\text{vac}\,|\,\text{not inf}}} &= OR \\
O_{\text{vac}\,|\,\text{inf}} &= OR\ O_{\text{vac}\,|\,\text{not inf}}
\end{aligned}$$

we can express $p_{\text{vac}}$ as a function of $p_{\text{inf}}$, $OR$ and $O_{\text{vac}\,|\,\text{inf}}$

$$\begin{aligned}
p_{\text{vac}} &= p_{\text{inf}}\ p_{\text{vac}\,|\,\text{inf}} + p_{\text{not inf}}\ p_{\text{vac}\,|\,\text{not inf}} \\
&= p_{\text{inf}} \frac{O_{\text{vac}\,|\,\text{inf}}}{1 + O_{\text{vac}\,|\,\text{inf}}} + p_{\text{not inf}} \frac{O_{\text{vac}\,|\,\text{not inf}}}{1 + O_{\text{vac}\,|\,\text{not inf}}} \\
&= p_{\text{inf}} \frac{\mathrm{OR}\ O_{\text{vac}\,|\,\text{not inf}}}{1 + \mathrm{OR}\ O_{\text{vac}\,|\,\text{not inf}}} + (1 - p_{\text{inf}}) \frac{O_{\text{vac}\,|\,\text{not inf}}}{1 + O_{\text{vac}\,|\,\text{not inf}}} \\
&= \frac{p_{\text{inf}}\ \mathrm{OR}\ O_{\text{vac}\,|\,\text{not inf}}}{1 + \mathrm{OR}\ O_{\text{vac}\,|\,\text{not inf}}} + \frac{O_{\text{vac}|\text{not inf}} - p_{\text{inf}}\ O_{\text{vac}\,|\,\text{not inf}}}{1 + O_{\text{vac}\,|\,\text{not inf}}}
\end{aligned}$$

For readability, let us define short hands for the above terms.

$$\begin{aligned}
p_{\text{vac}} &= v \\
p_{\text{inf}} &= i \\
O_{\text{vac}\,|\,\text{not inf}} &= d \text{ (for o}\mathbf{d}\text{ds)} \\
\mathrm{OR} &= r \text{ (for odds }\mathbf{r}\text{atio)}
\end{aligned}$$

Using the new labels, rearranging the terms, and equating to zero renders a quadratic equation of $d$ ($O_{\text{vac | not inf}}$):

$$\begin{aligned} v &= \frac{ird}{1+rd} + \frac{d-id}{1+d} \\ 0 &= v(1+rd)(1+d) \quad - ird(1+d) \quad - (d-id)(1+rd) \\ &= v + vrd + vd + vrd^2 - ird - ird^2 \quad - d + id - rd^2 + ird^2 \\ &= (vr-r)d^2 + (vr + v - ir + i - 1)d + v \end{aligned}$$

Making the following substitutions,

$$\begin{aligned} (vr - r) &= a \\ (vr + v - ir + i - 1) &= b \\ v &= c \end{aligned}$$

allows us to solve the quadratic formula, rendering a solution for $O_{\text{vac|not inf}}$, which allows us to compute the joint probability of being vaccinated and infected, and finally the probability of being immune.

$$\frac{-b \pm \sqrt{(b^2 + 4ac)}}{2a} = d = O_{\text{vac | not inf}}$$

$$O_{\text{vac | inf}} = O_{\text{vac | not inf}} \text{ OR}$$

$$p_{\text{vac | inf}} = \frac{O_{\text{vac | inf}}}{1 + O_{\text{vac | inf}}}$$

$$p_{\text{vac} \cap \text{inf}} = p_{\text{vac}}\, p_{\text{vac | inf}}$$

**Waning functions**

We propose four simplistic functions of waning of immunity (Figure S1), under three scenarios: a baseline scenario, used in the analyses in the manuscript, and an additional optimistic and pessimistic scenario (dotted lines in Figure S1) for sensitivity analyses.

Baseline scenario:

- Protection against infection conferred by vaccination or natural infection is 80% for the first two months after natural infection or vaccination, then declines to 50% in month 4, and then declines more slowly over the next five months, such that the protection nine months after infection or vaccination is 25% and constant thereafter.
- Protection against infection conferred by the combination of vaccination and natural infection is 90% and does not decline.
- Protection against severe outcomes conferred by vaccination or natural infection is 95% for the first six months after infection or complete vaccination, and then declines by 10 percent points every six months.
- Protection against severe outcomes conferred by the combination of natural infection and vaccination is 95% and does not decline.

Optimistic scenario:

- Protection against infection conferred by vaccination or natural infection is 90% for the first two months after natural infection or vaccination, then declines to 75% over the next two months, and then declines further over the next five months, such that the protection nine months after infection or vaccination is 50% and constant thereafter.
- Protection against infection conferred by the combination of vaccination and natural infection is 95% and does not decline.
- Protection against severe outcomes conferred by vaccination or natural infection is 100% for the first six months after the infection or complete vaccination, and then declines by 5 percent points every six months.
- Protection against severe outcomes accrued by the combination of natural infection and vaccination is 100% and does not decline.

Pessimistic scenario:

- Protection against infection conferred by vaccination or natural infection is 75% for the first two months after natural infection or vaccination, then declines to 45% over the next two months, and then declines further over the next five months, such that the protection nine months after infection or vaccination is 20% and constant thereafter.
- Protection against infection conferred by the combination of vaccination and natural infection is 80% for the first six months after being previously infected and vaccinated, and then declines by 10 percent points every six months.
- Protection against severe outcomes conferred by vaccination or natural infection is 100% for the first six months after the infection or complete vaccination, and then declines by 20 percent points every six months.
- Protection against severe outcomes conferred by the combination of natural infection and vaccination is 90% for the first six months after being previously infected and vaccinated, and then declines by 10 percent points every six months.

**Validation of Household Pulse Survey data**

We fit a logistic regression model on the Axios-Ipsos Coronavirus Survey data[6], similar to the logistic regression we used for the Household Pulse Survey data, that was used in the main analyses. The Axios-Ipsos survey data does not have state-specific estimates, rendering a simplified regression equation:

$$\begin{aligned} \text{logit}(\theta) &= \mathrm{b}_0 + X_{\text{week}}\mathbf{b}_{\text{week}} + X_{\text{covid}}\mathrm{b}_{\text{covid}} \\ \mathcal{L}(\theta|W,w) &= \theta^{w}(1-\theta)^{W-w} \\ b_{(\cdot)} &\sim \mathcal{N}(0,1000) \end{aligned}$$

**Table S1:** Means and standard deviations of the log-normal prior distributions for odds ratio of vaccination given infection status for each US state.

| State | Odds ratio | Lower bound 95% CrI | Upper bound 95% CrI |
|---|---|---|---|
| Florida | 0.400 | 0.361 | 0.444 |
| Michigan | 0.455 | 0.390 | 0.529 |
| Oregon | 0.469 | 0.397 | 0.555 |
| Washington | 0.47 | 0.401 | 0.551 |
| North Carolina | 0.475 | 0.424 | 0.532 |
| Pennsylvania | 0.484 | 0.428 | 0.547 |
| Alaska | 0.488 | 0.407 | 0.586 |
| Illinois | 0.495 | 0.441 | 0.557 |
| New Mexico | 0.501 | 0.403 | 0.623 |
| Maryland | 0.503 | 0.425 | 0.596 |
| New Jersey | 0.504 | 0.439 | 0.578 |
| Arizona | 0.505 | 0.444 | 0.574 |
| New York | 0.505 | 0.46 | 0.556 |
| Vermont | 0.507 | 0.406 | 0.633 |
| Hawaii | 0.508 | 0.417 | 0.618 |
| Delaware | 0.511 | 0.421 | 0.621 |
| Indiana | 0.512 | 0.449 | 0.584 |
| West Virginia | 0.512 | 0.431 | 0.608 |
| Connecticut | 0.516 | 0.44 | 0.605 |
| North Dakota | 0.516 | 0.431 | 0.617 |
| Idaho | 0.516 | 0.434 | 0.614 |
| Colorado | 0.516 | 0.447 | 0.596 |
| Nevada | 0.517 | 0.431 | 0.621 |
| Massachusetts | 0.518 | 0.441 | 0.607 |
| Ohio | 0.518 | 0.466 | 0.575 |
| Wyoming | 0.519 | 0.427 | 0.631 |
| Rhode Island | 0.519 | 0.432 | 0.625 |
| Kansas | 0.521 | 0.441 | 0.616 |
| Minnesota | 0.523 | 0.446 | 0.612 |
| Maine | 0.525 | 0.442 | 0.623 |
| Mississippi | 0.526 | 0.445 | 0.621 |
| District of Columbia | 0.53 | 0.438 | 0.641 |
| Georgia | 0.53 | 0.472 | 0.595 |
| Tennessee | 0.532 | 0.457 | 0.62 |
| New Hampshire | 0.534 | 0.435 | 0.656 |

| | | | |
|---|---|---|---|
| Montana | 0.535 | 0.439 | 0.651 |
| Oklahoma | 0.535 | 0.456 | 0.627 |
| Missouri | 0.535 | 0.467 | 0.614 |
| Utah | 0.536 | 0.451 | 0.637 |
| Nebraska | 0.537 | 0.435 | 0.663 |
| Virginia | 0.538 | 0.459 | 0.629 |
| South Dakota | 0.538 | 0.444 | 0.653 |
| Arkansas | 0.541 | 0.456 | 0.643 |
| South Carolina | 0.541 | 0.464 | 0.632 |
| California | 0.545 | 0.502 | 0.591 |
| Iowa | 0.554 | 0.472 | 0.652 |
| Wisconsin | 0.555 | 0.483 | 0.638 |
| Alabama | 0.56 | 0.484 | 0.649 |
| Louisiana | 0.567 | 0.487 | 0.659 |
| Kentucky | 0.569 | 0.486 | 0.665 |
| Texas | 0.581 | 0.532 | 0.634 |

**Table S2:** Relative risk of infection and severe outcomes (Omicron versus pre-Omicron) for those with protection.

| | RR of infection | RR of infection (boosted) | RR of severe disease | RR of severe disease (boosted) |
|---|---|---|---|---|
| High immune escape | .1 | .4 | .7 | .8 |
| Medium immune escape | .3 | .6 | .8 | .9 |
| Low immune escape | .5 | .8 | .9 | .95 |

**Table S3:** Effective protection outcomes for each US state on December 1, 2021 under base-case, pessimistic and optimistic scenarios.

| State | *Effective protection* against infection; base-case scenario (pessimistic; optimistic); no immune escape | *Effective protection* against infection with Omicron; pessimistic waning + high immune escape scenario (95%CrI) | *Effective protection* against infection with Omicron; base-case waning + medium immune escape scenario (95%CrI) | *Effective protection* against infection with Omicron; optimistic waning + low immune escape scenario (95%CrI) |
|---|---|---|---|---|
| Alabama | 52.8% (46.2-63.7) | 11.5% (11.2-11.9) | 20.7% (19.9-21.7) | 27.4% (26.2-28.6) |
| Alaska | 57.7% (50.8-66.5) | 14% (13.6-14.4) | 24.2% (23.2-25.4) | 31.4% (29.8-33.1) |
| Arizona | 58.8% (51.2-68.5) | 12.4% (12.1-12.7) | 22.9% (22.1-23.8) | 30.1% (29-31.2) |
| Arkansas | 50.5% (44.1-61.2) | 11.9% (11.6-12.4) | 20.6% (19.8-21.9) | 26.8% (25.4-28.5) |
| California | 55.3% (47.9-65.4) | 12.4% (12-12.8) | 22.1% (20.9-23.8) | 28.7% (27-31.1) |
| Colorado | 63.2% (55-71.8) | 15% (14.8-15.4) | 26.4% (25.3-27.8) | 33.9% (32.3-35.9) |
| Connecticut | 58.8% (50.8-69.1) | 13.7% (13.5-14.2) | 24.1% (23-25.9) | 31% (29.3-33.6) |
| Delaware | 56.8% (49.4-66.6) | 13.2% (12.9-13.6) | 23.2% (22.2-24.7) | 30% (28.4-32.2) |
| District of Columbia | 55.7% (48.3-66.4) | 11.4% (11.1-11.8) | 21.2% (20-23) | 28% (26.3-30.7) |
| Florida | 60.5% (53-70.1) | 12.6% (12.3-13.1) | 23.5% (22.3-24.8) | 31.2% (29.5-33.1) |
| Georgia | 51.8% (45.2-62.7) | 11.1% (10.8-11.6) | 20.2% (19.2-21.2) | 26.6% (25.3-28) |
| Hawaii | 46.4% (40.2-60.4) | 10.1% (9.9-10.5) | 17.8% (16.9-19.6) | 23.4% (22.1-26.2) |

| | | | | |
|---|---|---|---|---|
| Idaho | 54.7% (48.1-64.2) | 13.8% (13.5-14.2) | 23.4% (22.6-24.3) | 30.1% (28.8-31.4) |
| Illinois | 52.6% (45.3-62.9) | 13.1% (12.8-13.6) | 22.2% (21.1-23.9) | 28.2% (26.6-30.6) |
| Indiana | 47.9% (41.6-58.3) | 11.6% (11.3-12.2) | 19.8% (18.8-21.4) | 25.5% (24-27.7) |
| Iowa | 53% (45.8-62.4) | 14.7% (14.5-15.2) | 23.9% (23.1-25.2) | 29.8% (28.5-31.7) |
| Kansas | 52.6% (45.7-62.5) | 12.5% (12.2-12.9) | 21.7% (20.7-23.1) | 27.9% (26.5-30) |
| Kentucky | 55.1% (48.2-64.8) | 13.3% (13-13.7) | 23% (22.1-24.1) | 29.7% (28.3-31.2) |
| Louisiana | 51.1% (44.4-62) | 11.6% (11.3-12.1) | 20.5% (19.6-21.7) | 26.7% (25.3-28.3) |
| Maine | 55.5% (48.3-66.2) | 14.6% (14.4-15) | 24.2% (23.3-25.9) | 30.7% (29.2-33.3) |
| Maryland | 55.1% (47.8-65.4) | 13.4% (13.2-13.9) | 23.1% (22-24.8) | 29.5% (28-32.1) |
| Massachusetts | 57.4% (49.6-67.7) | 13.3% (13-13.8) | 23.4% (22.3-25.2) | 30.1% (28.4-32.7) |
| Michigan | 53.3% (46.3-63.2) | 14% (13.7-14.5) | 23.2% (22.4-24.5) | 29.4% (28.1-31.1) |
| Minnesota | 54.2% (46.9-63.3) | 15.1% (14.8-15.5) | 24.5% (23.7-25.9) | 30.6% (29.2-32.7) |
| Mississippi | 50.8% (44.2-61.9) | 11.2% (10.9-11.7) | 20% (19.1-21.2) | 26.2% (24.9-27.8) |
| Missouri | 46.8% (40.6-57.2) | 11.7% (11.4-12.3) | 19.7% (18.8-21.3) | 25.2% (23.8-27.5) |
| Montana | 55.1% (48.1-64.4) | 14.4% (14.1-14.9) | 24.1% (23.2-25.2) | 30.6% (29.3-32.2) |

| | | | | |
|---|---|---|---|---|
| Nebraska | 51.8% (45.2-61.3) | 13.4% (13-14.1) | 22.4% (21.1-24.3) | 28.6% (26.4-31.4) |
| Nevada | 55.4% (48.3-65.5) | 11.7% (11.1-12.4) | 21.5% (19.7-23.6) | 28.4% (25.7-31.5) |
| New Hampshire | 54.9% (48.6-67.3) | 8.4% (8.1-8.9) | 18.1% (16.9-20.1) | 25.7% (23.9-28.7) |
| New Jersey | 57.6% (49.8-67.4) | 12.6% (12.3-13) | 22.8% (21.6-24.5) | 29.7% (27.9-32.1) |
| New Mexico | 62.4% (54.4-71.1) | 14.7% (14.5-15) | 25.9% (25.2-26.7) | 33.4% (32.4-34.4) |
| New York | 57.8% (50.1-67.9) | 11.1% (10.8-11.5) | 21.4% (20.1-23.2) | 28.6% (26.7-31) |
| North Carolina | 46.2% (40.1-57.4) | 9.4% (8.9-10.1) | 17.3% (15.8-19.6) | 23% (20.8-26.5) |
| North Dakota | 53.2% (46-63.1) | 13.6% (13.3-14) | 22.8% (22.1-23.8) | 28.9% (27.9-30.1) |
| Ohio | 48.3% (41.8-58.2) | 13% (12.7-13.7) | 21.3% (20.3-23.2) | 26.7% (25.2-29.6) |
| Oklahoma | 57.8% (50.8-67.4) | 12.3% (12-12.6) | 22.6% (21.8-23.5) | 29.9% (28.7-31.1) |
| Oregon | 52.8% (46.2-63) | 13.2% (13-13.8) | 22.4% (21.3-24.5) | 28.8% (26.9-32.1) |
| Pennsylvania | 56.3% (49.1-66.8) | 10.8% (10.4-11.2) | 20.7% (19.4-22.6) | 27.9% (26-30.6) |
| Rhode Island | 61.2% (53.2-70.6) | 14.3% (14-14.6) | 25.2% (24.2-26.6) | 32.5% (31-34.4) |
| South Carolina | 51.9% (45.2-62.3) | 12% (11.7-12.5) | 21% (20-22.3) | 27.3% (25.9-29.2) |
| South Dakota | 55.4% (48.4-64.7) | 13.1% (12.9-13.5) | 22.9% (22.1-24.1) | 29.6% (28.5-31.2) |

| | | | | |
|---|---|---|---|---|
| Tennessee | 51.7% (44.9-62.1) | 12.7% (12.3-13.5) | 21.6% (20.4-23.7) | 27.8% (25.9-30.8) |
| Texas | 54.4% (47.3-64.3) | 11.5% (11.2-11.9) | 21.1% (20.1-22.4) | 27.8% (26.3-29.6) |
| Utah | 49% (42.9-59) | 11.2% (10.8-11.7) | 19.7% (18.6-21) | 25.8% (24.2-27.7) |
| Vermont | 50.4% (43.6-59.2) | 15.5% (15.4-15.9) | 24.2% (23.5-25.7) | 29.6% (28.4-32.1) |
| Virginia | 50.1% (43.4-60.2) | 12.6% (12.3-13.1) | 21.2% (20.3-22.9) | 27.1% (25.5-29.6) |
| Washington | 50.9% (44.3-61.5) | 12.8% (12.5-13.3) | 21.6% (20.6-23.3) | 27.6% (26-30.3) |
| West Virginia | 41.6% (36.8-53.1) | 7.3% (6.8-8) | 14.4% (13.2-15.8) | 20.1% (18.4-22) |
| Wisconsin | 52.9% (45.7-62.4) | 14.3% (14.1-14.9) | 23.5% (22.7-25.3) | 29.5% (28.1-32.1) |
| Wyoming | 56.2% (49.4-66) | 13.8% (13.3-14.7) | 23.7% (22.1-25.8) | 30.7% (28.2-33.8) |

**Table S4:** Effective protection against severe disease for each US state on December 1, 2021 under base-case, pessimistic and optimistic scenarios.

| State | *Effective protection* against severe disease; base-case scenario (pessimistic; optimistic); no immune escape | *Effective protection* against severe disease with Omicron; pessimistic waning + high immune escape scenario (95%CrI) | *Effective protection* against severe disease with Omicron; base-case waning + medium immune escape scenario (95%CrI) | *Effective protection* against severe disease with Omicron; optimistic waning + low immune escape scenario (95%CrI) |
|---|---|---|---|---|
| Alabama | 74.1% (67.2-79.2) | 57.3% (54.3-60.5) | 61% (57.9-64.3) | 61.3% (58.5-64.2) |
| Alaska | 73.9% (68.2-78.5) | 57.5% (54.9-60.5) | 61.6% (58.8-64.8) | 62.5% (59.5-65.7) |
| Arizona | 77.3% (70.1-82.5) | 59.7% (57.9-61.5) | 63.7% (61.9-65.5) | 64% (62.4-65.4) |
| Arkansas | 71.5% (64.8-76.4) | 55.5% (52.4-59.3) | 59.1% (55.8-63) | 59.2% (56-62.8) |
| California | 75.3% (68.8-80.1) | 58.1% (56.7-60) | 62.2% (60.7-64.3) | 62.8% (61.2-64.9) |
| Colorado | 78.9% (72.6-83.6) | 61.3% (60.6-62.2) | 65.8% (64.9-66.7) | 66.5% (65.6-67.6) |
| Connecticut | 79.5% (72.8-84.4) | 61.5% (61-62.3) | 65.9% (65.3-66.8) | 66.6% (65.9-67.6) |
| Delaware | 75.8% (69.2-80.7) | 58.8% (57.1-60.9) | 62.8% (61-65.1) | 63.2% (61.5-65.4) |
| District of Columbia | 77.3% (70.7-82.3) | 59.2% (58.4-60.5) | 63.4% (62.5-64.8) | 64.4% (63.5-65.8) |
| Florida | 78.5% (72.3-83.3) | 60.3% (58.8-62) | 64.7% (63-66.5) | 66% (64.3-67.7) |
| Georgia | 73.4% (66.3-78.6) | 56.7% (53.7-60.1) | 60.3% (57.2-63.7) | 60.4% (57.6-63.4) |
| Hawaii | 77.3% (71-82.3) | 59% (58.8-59.6) | 63.2% (62.9-64) | 64.5% (64-65.6) |

| | | | | |
|---|---|---|---|---|
| Idaho | 72.5% (66.3-77.2) | 56.6% (53.3-60.4) | 60.4% (57-64.3) | 60.8% (57.4-64.5) |
| Illinois | 73.3% (66.3-78.3) | 57.2% (55.4-59.6) | 60.9% (59.1-63.5) | 60.7% (59-63.3) |
| Indiana | 68.8% (62-73.7) | 53.6% (50.6-57.7) | 57% (53.8-61.3) | 56.7% (53.7-60.8) |
| Iowa | 71.4% (64.5-76.2) | 56.3% (53.9-59.6) | 60% (57.4-63.4) | 59.4% (56.9-62.7) |
| Kansas | 72.3% (65.8-77) | 56.1% (53.7-59.1) | 59.9% (57.4-63.1) | 60.2% (57.7-63.3) |
| Kentucky | 73.9% (67.7-78.6) | 57.4% (54.6-60.7) | 61.4% (58.4-64.8) | 62% (59-65.2) |
| Louisiana | 72% (64.8-77.3) | 56% (53.1-59.4) | 59.5% (56.5-62.8) | 59.2% (56.5-62.1) |
| Maine | 77.2% (71.1-82) | 60.2% (59.3-61.7) | 64.4% (63.4-66.2) | 65.2% (64-67.3) |
| Maryland | 75.8% (69.4-80.6) | 58.8% (57.8-60.3) | 62.9% (61.8-64.6) | 63.5% (62.3-65.3) |
| Massachusetts | 77.9% (71-82.9) | 60.3% (59.3-61.7) | 64.5% (63.4-66.1) | 64.9% (63.8-66.5) |
| Michigan | 72.6% (65.5-77.7) | 57.1% (54.2-60.6) | 60.7% (57.7-64.2) | 60.2% (57.4-63.4) |
| Minnesota | 71.9% (65.4-76.6) | 56.7% (54.6-59.6) | 60.4% (58.3-63.5) | 60.2% (58-63.2) |
| Mississippi | 72.4% (65.2-77.7) | 56.1% (53.2-59.5) | 59.6% (56.6-63.1) | 59.4% (56.7-62.5) |
| Missouri | 67.8% (61.4-72.5) | 52.8% (49.7-57.3) | 56.2% (52.9-60.9) | 56.1% (52.9-60.7) |
| Montana | 72.9% (66.6-77.7) | 57.2% (54.3-60.7) | 61% (57.9-64.7) | 61.2% (58.1-64.8) |

| | | | | |
|---|---|---|---|---|
| Nebraska | 70.3% (63.8-75) | 55.1% (51.1-59.6) | 58.7% (54.4-63.4) | 58.6% (54.3-63.4) |
| Nevada | 75% (68.5-80) | 57.8% (54.3-61.2) | 61.7% (58-65.5) | 62.4% (58.6-66.3) |
| New Hampshire | 80.2% (74.8-84.9) | 60% (59.5-60.9) | 64.7% (64.1-65.7) | 67.6% (67-68.7) |
| New Jersey | 76.8% (69.8-81.8) | 59.3% (58.2-60.7) | 63.4% (62.2-64.9) | 63.7% (62.6-65.2) |
| New Mexico | 78.4% (71.8-83.3) | 61% (59.6-62.4) | 65.2% (63.8-66.6) | 65.8% (64.5-66.9) |
| New York | 77.4% (70.5-82.5) | 59.2% (58.2-60.6) | 63.4% (62.2-64.8) | 64.1% (63-65.4) |
| North Carolina | 69.4% (62.8-74.2) | 53.3% (50-57.7) | 56.7% (53.3-61.5) | 57.1% (53.5-62.1) |
| North Dakota | 72.6% (64.9-77.8) | 57% (54.2-60.2) | 60.5% (57.6-63.7) | 59.6% (56.9-62.5) |
| Ohio | 68.2% (61.5-72.9) | 53.5% (50.6-58.3) | 56.9% (53.8-62) | 56.4% (53.3-61.6) |
| Oklahoma | 76% (69.8-80.8) | 58.5% (56.3-60.8) | 62.7% (60.3-65.1) | 63.7% (61.4-65.9) |
| Oregon | 73.1% (67.4-77.7) | 56.8% (55-59.7) | 60.8% (58.8-64) | 61.7% (59.5-65.3) |
| Pennsylvania | 77.3% (70.9-82.2) | 58.9% (57.8-60.4) | 63.2% (62-64.9) | 64.5% (63.2-66.2) |
| Rhode Island | 78.7% (71.9-83.7) | 61.1% (59.9-62.5) | 65.3% (64.1-66.9) | 65.8% (64.6-67.2) |
| South Carolina | 72.2% (65.5-77.2) | 56% (53.1-59.6) | 59.7% (56.7-63.4) | 59.8% (56.9-63.3) |
| South Dakota | 73.3% (66.7-78.2) | 57.1% (55-59.5) | 60.9% (58.8-63.4) | 61.1% (59.1-63.5) |

| | | | | |
|---|---|---|---|---|
| Tennessee | 72.3% (65.4-77.3) | 56.4% (52.8-61.4) | 60% (56.3-65.4) | 59.8% (56.1-65.3) |
| Texas | 73.6% (67-78.5) | 56.8% (54.8-59.1) | 60.6% (58.5-63) | 61.1% (59.1-63.3) |
| Utah | 68.8% (62.8-73.4) | 53.2% (50-57.2) | 56.8% (53.4-61) | 57.4% (54-61.4) |
| Vermont | 67.8% (62-72) | 53.7% (51.7-57.4) | 57.4% (55.2-61.4) | 57.2% (54.8-61.6) |
| Virginia | 70.7% (64.4-75.3) | 55% (53-58.2) | 58.7% (56.6-62.1) | 59% (56.8-62.5) |
| Washington | 72.3% (66.4-76.8) | 56.1% (54.5-58.6) | 60% (58.3-62.8) | 60.8% (58.9-63.7) |
| West Virginia | 65.4% (59.2-70.1) | 49.8% (44.3-56.5) | 53% (47.3-60) | 53.6% (48.1-60.4) |
| Wisconsin | 71.7% (65.1-76.4) | 56.3% (54.2-59.9) | 60.1% (57.8-63.8) | 59.9% (57.5-63.8) |
| Wyoming | 74.7% (68.3-79.5) | 58.2% (53.6-63.3) | 62.1% (57.2-67.5) | 62.6% (57.4-68.2) |

**Figure S1:** Assumed waning curves for protection against infection and severe disease, from natural infection and/or vaccination.

Footnote: Dashed lines indicate the optimistic and pessimistic scenario.

**Figure S2:** County-level estimates of the fraction of the population with *immunological exposure* at four time-points between December 31, 2020 and December 1, 2021.

**Figure S3:** County-level estimates of the fraction of the population with *effective protection* against infection on December 1, 2021, assuming no, low, medium, or high immune escape.

**Figure S4:** Contribution of prior infection and vaccination to the fraction of the population *immunologically exposed* for each US state over time.

Footnote: Purple: percent ever infected and not vaccinated. Green: percent vaccinated and not prior infected. Blended color: percent ever infected and vaccinated. Darker shades: received booster.

**Figure S5:** Contribution of effective protection from prior infection and vaccination or both to the total effective protection for each US state over time.

Footnote: Purple: effective protection from those infected and not vaccinated. Green: effective protection from those vaccinated and not prior infected. Blended color: effective protection from those infected and vaccinated. Darker shades: received booster.

**Figure S6:** Comparison of immunity estimates for each US state, with credible intervals, with blood donor estimates of immunity[7].

Footnote: red lines indicate the blood donor estimates
