## Supplementary figures and images for "Population immunity to pre-Omicron and Omicron SARS-CoV-2 variants in US states and counties through December 1, 2021"

### supplementary-figures

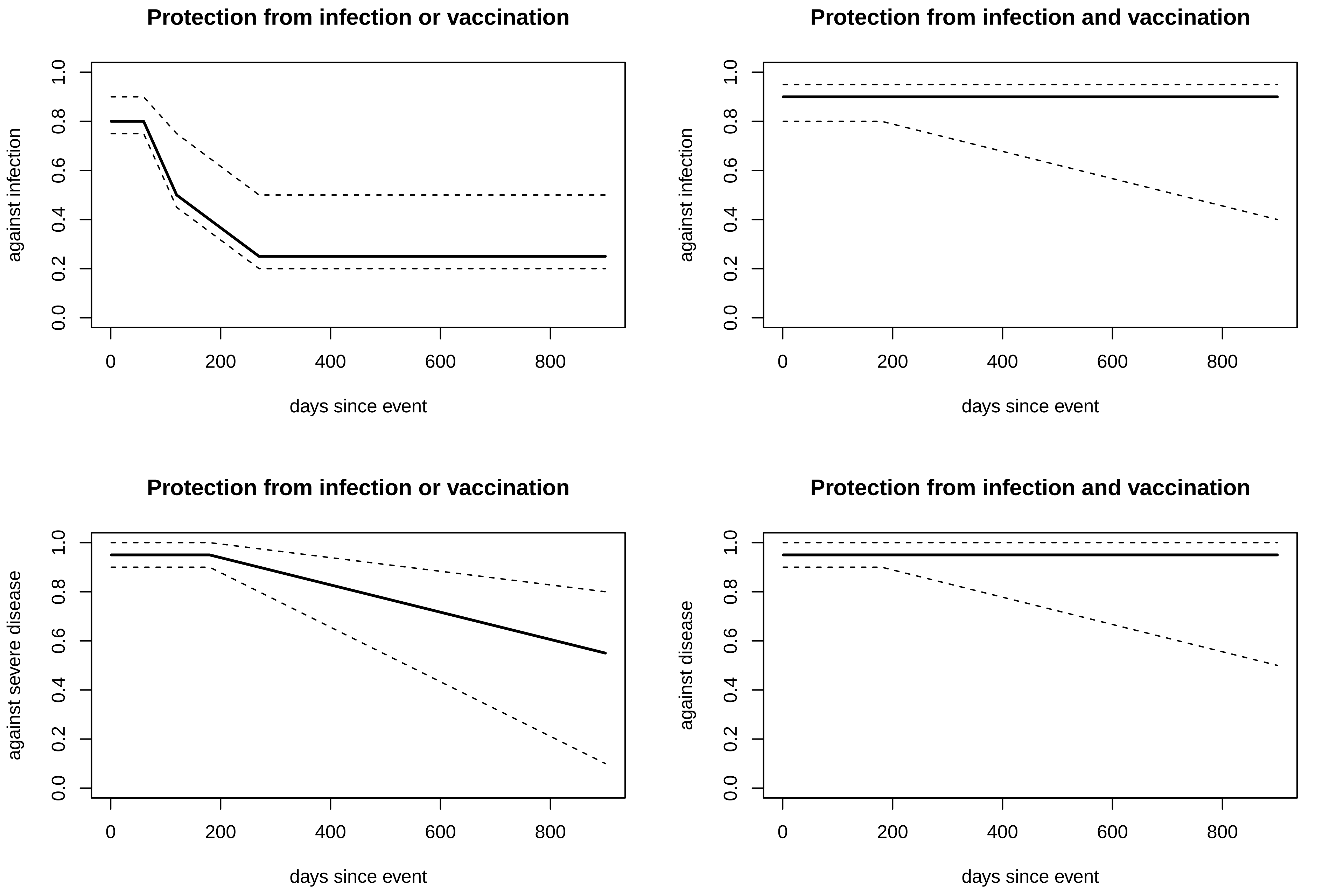

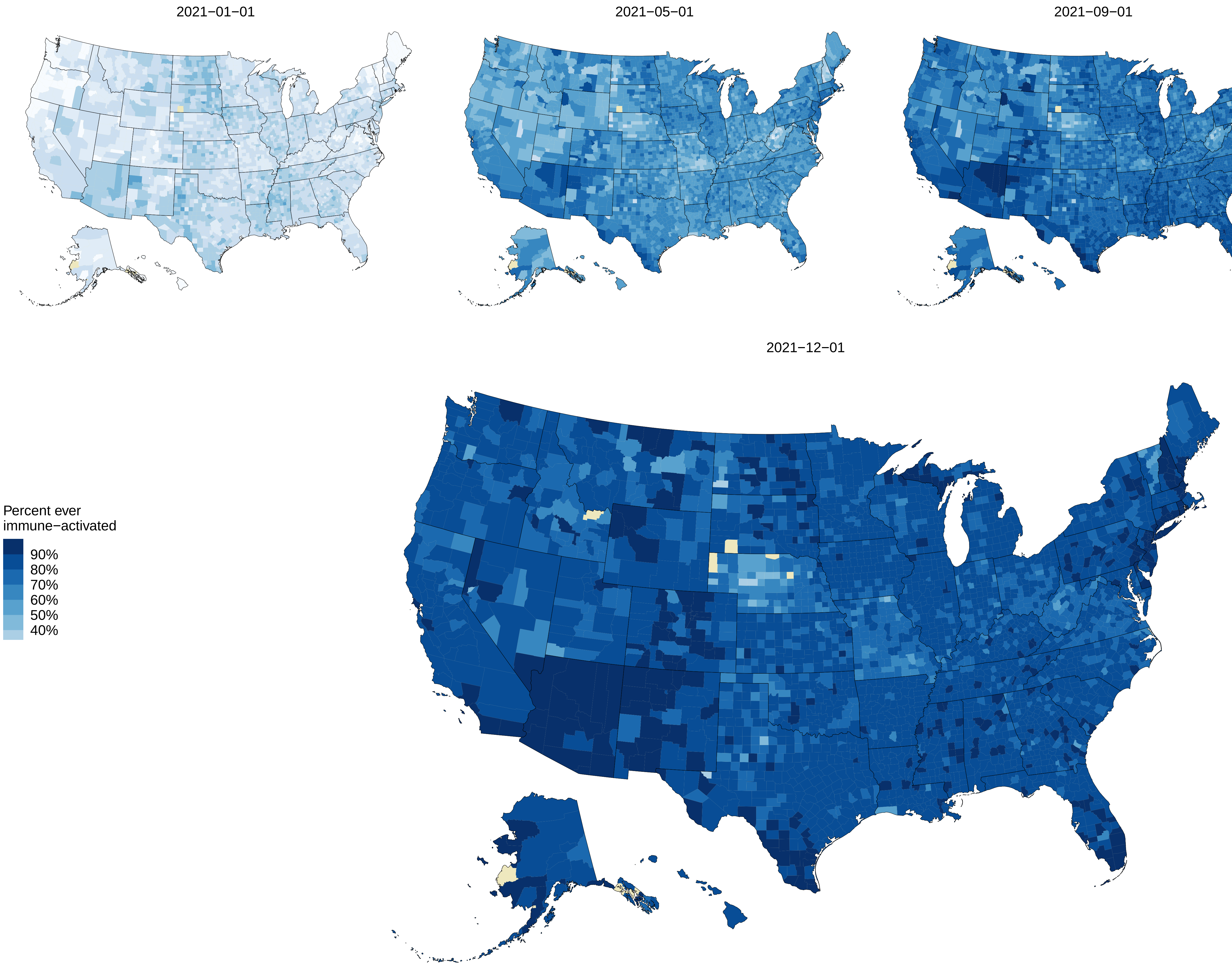

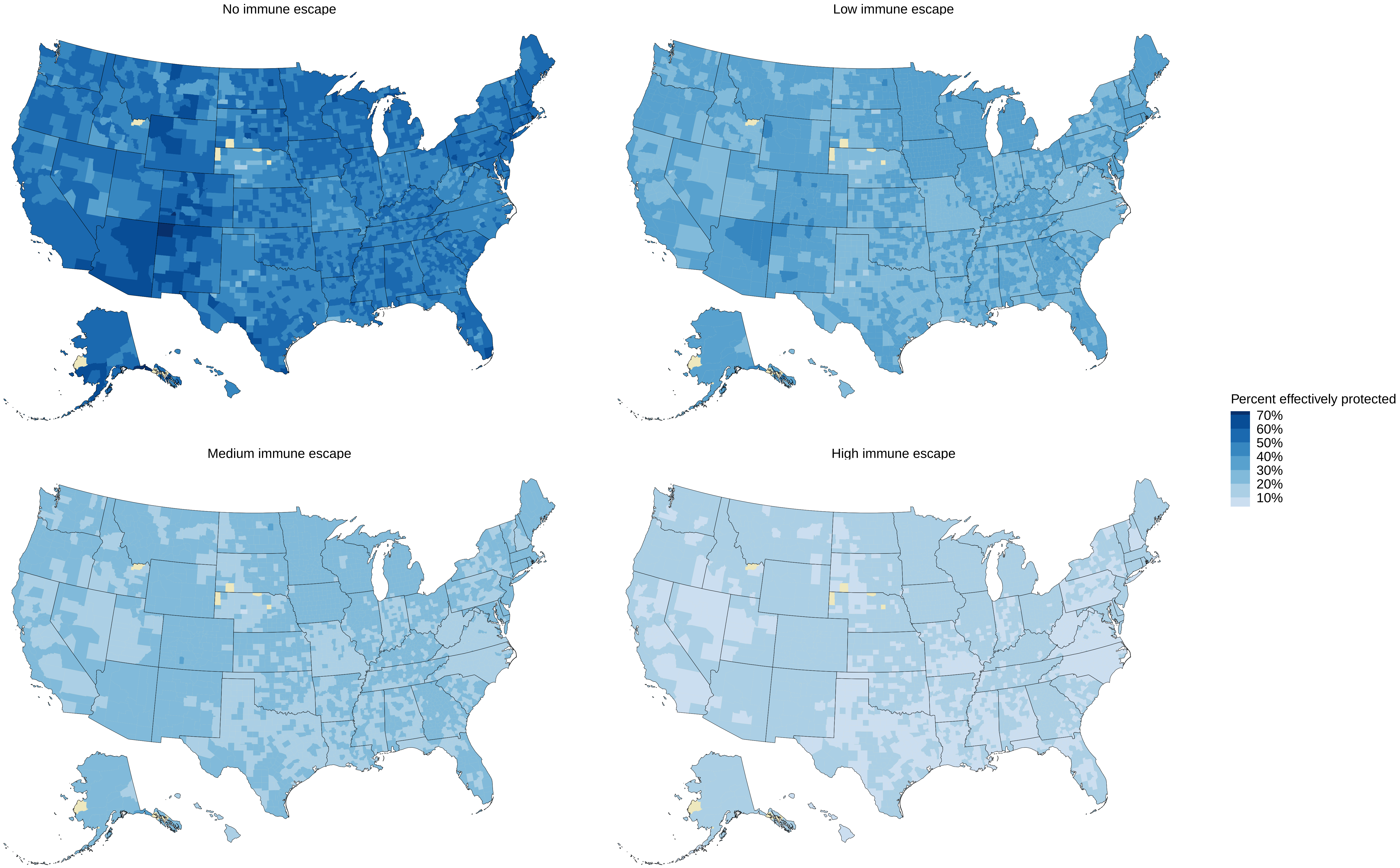

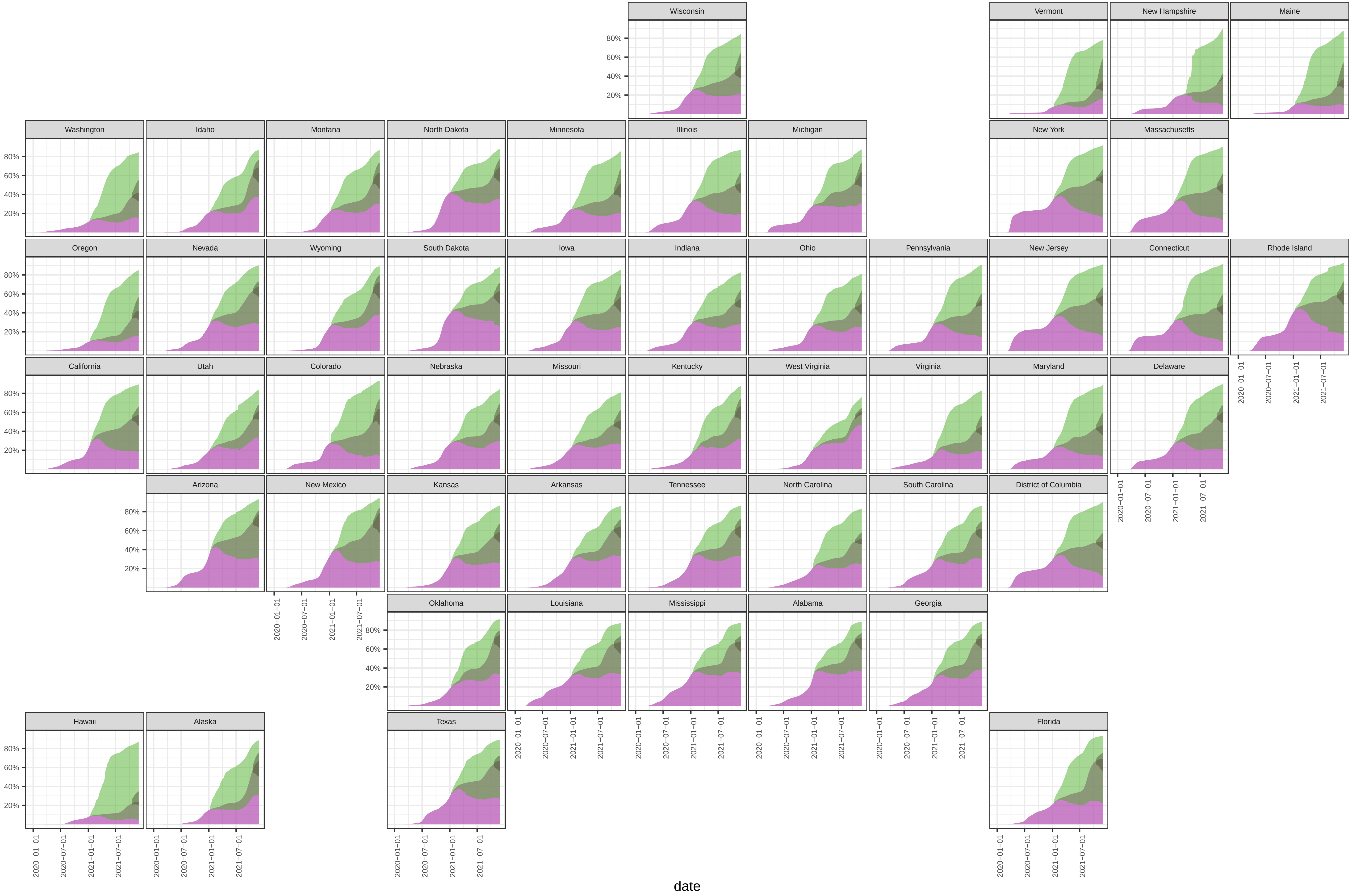

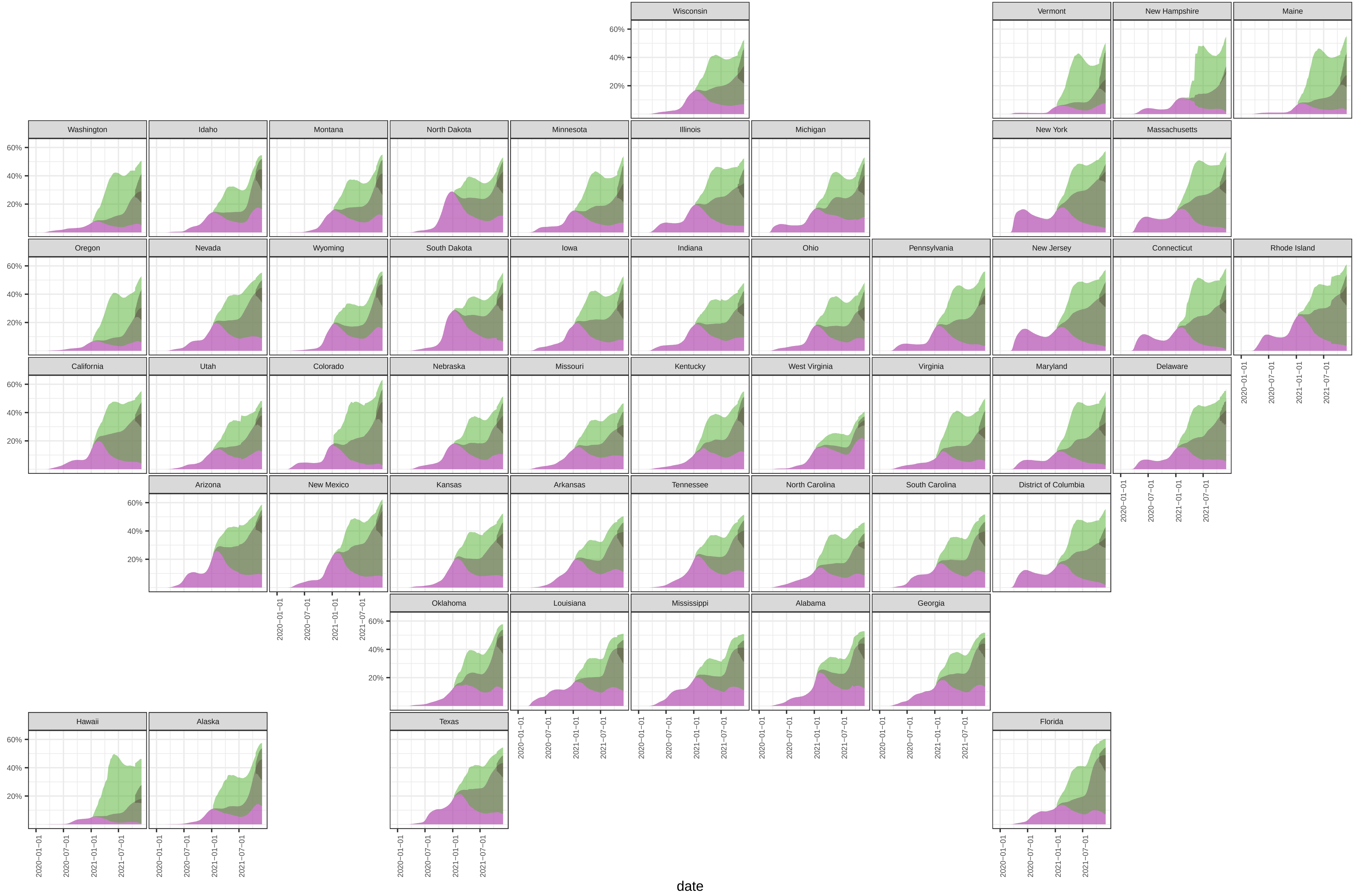

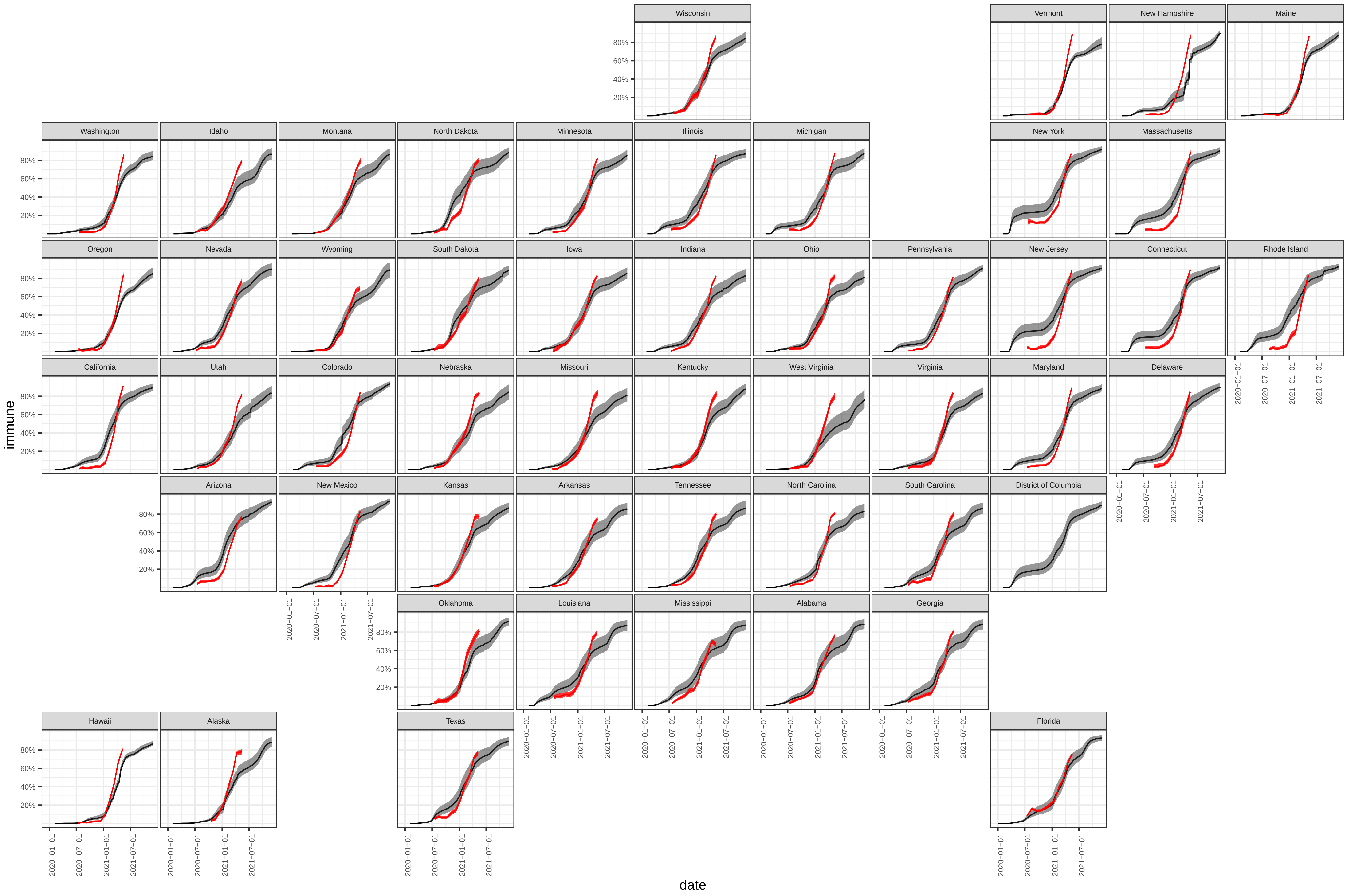
